# *Plasmodium knowlesi* infection is associated with elevated circulating biomarkers of brain injury and endothelial activation

**DOI:** 10.1101/2024.04.25.24306382

**Authors:** Cesc Bertran-Cobo, Elin Dumont, Naqib Rafieqin Noordin, Meng-Yee Lai, William Stone, Kevin KA Tetteh, Chris Drakeley, Sanjeev Krishna, Yee-Ling Lau, Samuel C Wassmer

## Abstract

**Introduction:** Malaria remains a major public health concern with substantial morbidity and mortality worldwide. In Malaysia, the emergence of *Plasmodium knowlesi* has led to a surge in zoonotic malaria cases and deaths in recent years. Signs of cerebral involvement have been observed in a non-comatose, fatal case of severe knowlesi infection, but the potential impact of this malaria species on the brain remains underexplored. To address this gap, we investigated circulating levels of brain injury, inflammation, and vascular biomarkers in a cohort of knowlesi-infected patients and controls.

**Methods:** Archived plasma samples from 19 patients with confirmed symptomatic knowlesi infection and 19 healthy, age-matched controls from Peninsular Malaysia were analysed. A total of 52 plasma biomarkers of brain injury, inflammation, and vascular activation were measured using Luminex and SIMOA assays. Wilcoxon tests were used to examine group differences, and biomarker profiles were explored through hierarchical clustering heatmap analysis.

**Results:** Bonferroni-corrected analyses revealed significantly elevated brain injury biomarker levels in knowlesi-infected patients, including S100B (p<0.0001), Tau (p=0.0007), UCH-L1 (p<0.0001), αSyn (p<0.0001), Park7 (p=0.0006), NRGN (p=0.0022), and TDP-43 (p=0.005). Compared to controls, levels were lower in the infected group for BDNF (p<0.0001), CaBD (p<0.0001), CNTN1 (p<0.0001), NCAM-1 (p<0.0001), GFAP (p=0.0013), and KLK6 (p=0.0126). Hierarchical clustering revealed distinct group profiles for circulating levels of brain injury and vascular activation biomarkers.

**Conclusions:** Our findings highlight for the first time the impact of *Plasmodium knowlesi* infection on the brain, with distinct alterations in cerebral injury and endothelial activation biomarker profiles compared to healthy controls. Further studies are warranted to investigate the pathophysiology and clinical significance of these altered surrogate markers, through both neuroimaging and long-term neurocognitive assessments.

## 1 Introduction

Malaria is a life-threatening, vector-borne infection caused by parasites of the genus *Plasmodium* that led to an estimated 249 million cases worldwide and approximately 608,000 deaths in 2022 (1). Malaria is endemic in Southeast Asia, where it poses a significant public health challenge. In Malaysia, *P. falciparum* and *P. vivax* have historically been the species responsible for most malaria infections and deaths, but thanks to national efforts in eradicating the disease, no indigenous cases have been reported since 2018 (1). However, an emerging concern in the country is *P. knowlesi* (*Pk*), a zoonotic species that has become a significant contributor to malaria infections in humans locally (2,3), with a total of 19,625 *Pk* cases and 57 deaths reported in Malaysia since 2017, including 2,500 cases and 9 deaths in 2022 alone (1). Malaysia accounts for most *Pk* infections globally, and this species is currently the major cause of human malaria in the country (4–6).

One of the most severe manifestations of *Plasmodium falciparum* infection is cerebral malaria, a life-threatening neurological complication characterized by coma (7) and pathological hallmarks such as petechial hemorrhages and parasite sequestration in the brain vasculature (8,9). Neurocognitive sequelae are frequent in survivors and can persist long after the infection has been treated (10,11). While cerebral malaria is predominantly associated with *P. falciparum* (7), recent case reports have highlighted instances of cerebral involvement and severe neurological complications in patients infected with other *Plasmodium* species (12–18). A high proportion of severe *Pk* infections has been reported in Southeast Asia (5,19), as well as fatal cases (18–20). A recent study also showed that, similarly to *P. falciparum*, *Pk*-infected erythrocytes are able bind to endothelial cells (21). Post-mortem findings in one fatal case of severe *Pk* malaria revealed brain pathology features similar to those seen in fatal falciparum cerebral malaria, including *Pk*-infected erythrocytes sequestration in the microvasculature (18), suggesting that *Pk* malaria may also affect the brain. Remarkably, coma was not observed in this patient (18), which contrasts with the World Health Organization (WHO) definition of cerebral malaria caused by *P. falciparum* (7).

We recently reported that patients with severe and uncomplicated falciparum malaria —and therefore without coma— have a wide range of brain changes detected using magnetic resonance imaging (MRI) (22,23). Severe malaria patients from our cohort had elevated plasma levels of the neurotrophic factor S100 calcium-binding protein β (S100B), a biomarker associated with central nervous system insults (24,25), which correlated with brain MRI features typically found in cerebral malaria (22). Overall, our findings suggest that there is a frequent and unrecognized impact of malaria infection on the brain in *falciparum* malaria, irrespective of coma. These effects are likely aggravated by acute kidney injury (AKI), with potential long-lasting repercussions on quality of life and productivity in survivors (26).

Despite the similar pathologies reported between cerebral malaria and fatal *Pk* infection (18) and the high incidence of kidney dysfunction in severe *Pk* malaria (27), the potential impact of *Pk* infection on the human brain remains largely unexplored. To bridge this important knowledge gap, we quantified plasma biomarkers of brain injury, vascular activation, and inflammation in a cohort of Malaysian *Pk*-infected patients and healthy controls.

## 2 Methods

### 2.1 Participants and samples

This study leveraged archived plasma samples from patients aged 18 years and above with malaria symptoms attending either a government hospital or private clinic in Johor, Selangor, Pahang, Perak, and Trengganu states between December 2019 and January 2023 (**Figure 1**). Recruitment of community-matched, age-matched uninfected controls was conducted via active screening of individuals from communities in Johor, Selangor, Negeri Sembilan, and Kedah. Subjects were approached for participation if they had no fever, were aged 18 years and above, and were part of any of the considered high-risk groups. A high-risk group is defined as individuals working in proximity with forest and forest fringes, i.e., servicemen, farmers, hunters, and natural resource collectors (2,28).

**Figure 1.**
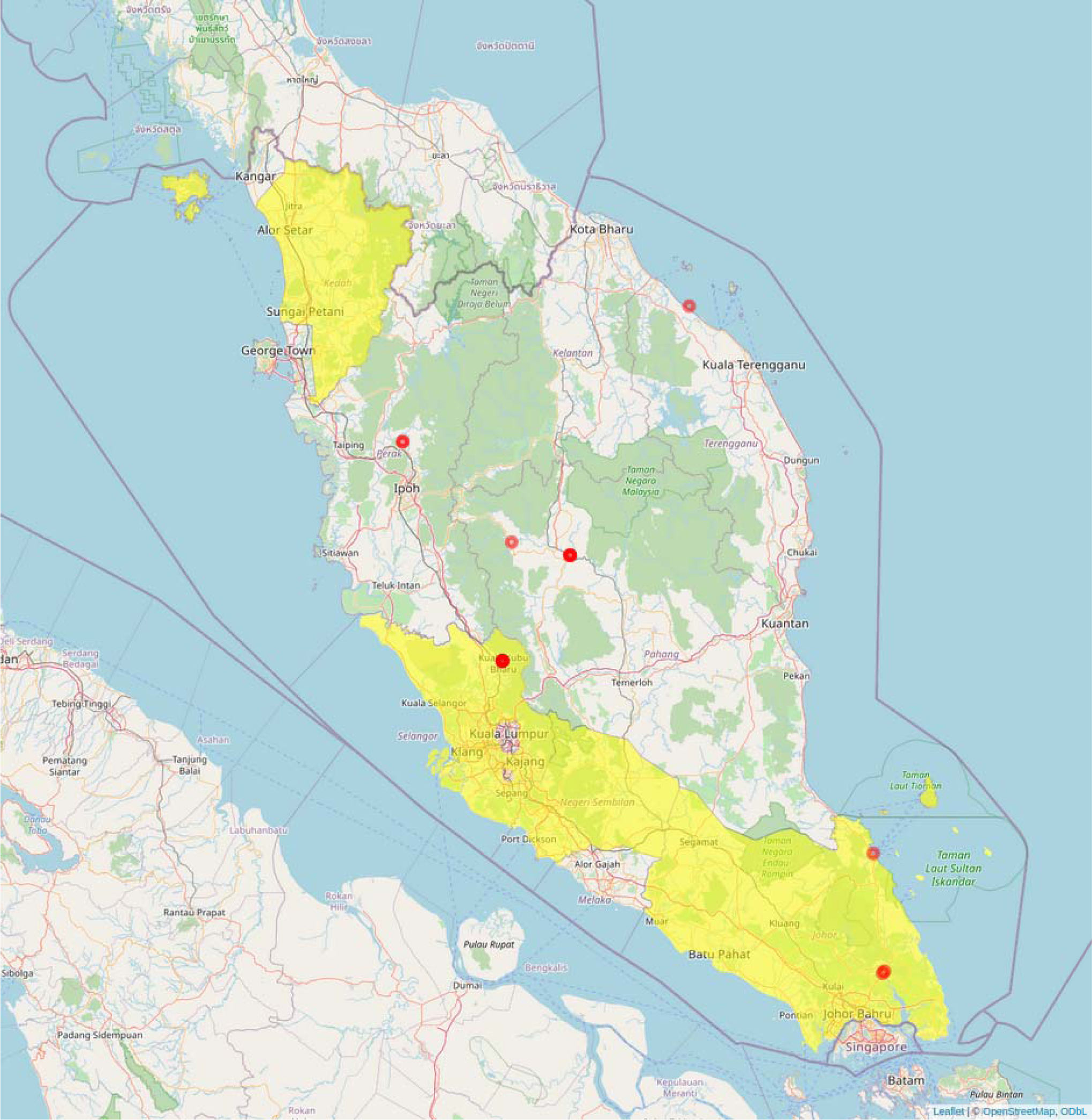
<u>Sample collection sites.</u>. Samples from *Pk*-infected patients were collected from government hospitals and private clinics in Johor, Selangor, Pahang, Perak, and Trengganu states (red). Samples from community-matched, age-matched uninfected controls were collected via active sample screening of communities in Johor, Selangor, Negeri Sembilan, and Kedah (yellow). Map was created in R with RStudio using Leaflet (https://leafletjs.com/) and OpenStreetMap (https://www.openstreetmap.org). An interactive version of the map can be found in GitHub (https://github.com/Cescualito/LSHTM_Wassmer_Pknowlesi_Malaysia).

Data collection procedures are described elsewhere (29). Briefly, samples were acquired using EDTA anti-coagulant tubes. Informed consent was obtained prior to sample collection. All participants were allowed sufficient time to consider their participation in the project. Individual data were collected and recorded anonymously. *Plasmodium* infection and speciation in each collected sample were confirmed by microscopic examination of Giemsa-stained blood smears and nested polymerase chain reaction (PCR) based on the 18S rRNA gene. Out of the 50 individuals sampled as part of the parent project, 38 had associated clinical information needed for this study, including parasitemia. The de-identified Malaysian samples, consisting of 19 infected patients and 19 healthy controls, were analysed.

### 2.2 Biomarker panel selection

A total of 52 plasma biomarkers were selected, including 21 markers of brain alterations or cerebral injury, 21 markers of infection and immune activation, and 10 vascular biomarkers. Our literature review-based selection followed the assumption that any plasma biomarker ever reported to be altered in any species of human malaria could potentially be altered in our cohort of *Pk* patients. For brain injury biomarkers, a second premise applied: since cerebral malaria can cause long-lasting cognitive disorders (10,11), any plasma marker reported to be increased or decreased in cognitively impaired subjects (e.g., neurodegenerative diseases, traumatic brain injury, etc) could also be altered in our patients.

To inform the biomarker panel selection, a systematic search of scientific literature on biomarker plasma levels in human patients was conducted in PubMed database, including all relevant publications from 2010 onwards. The results of this systematic search are comprehensively summarized in **Table 1** (brain injury biomarkers), **Supplementary Table 1** (infection and immune activation biomarkers) and **Supplementary Table 2** (vascular biomarkers).

**Table 1.**
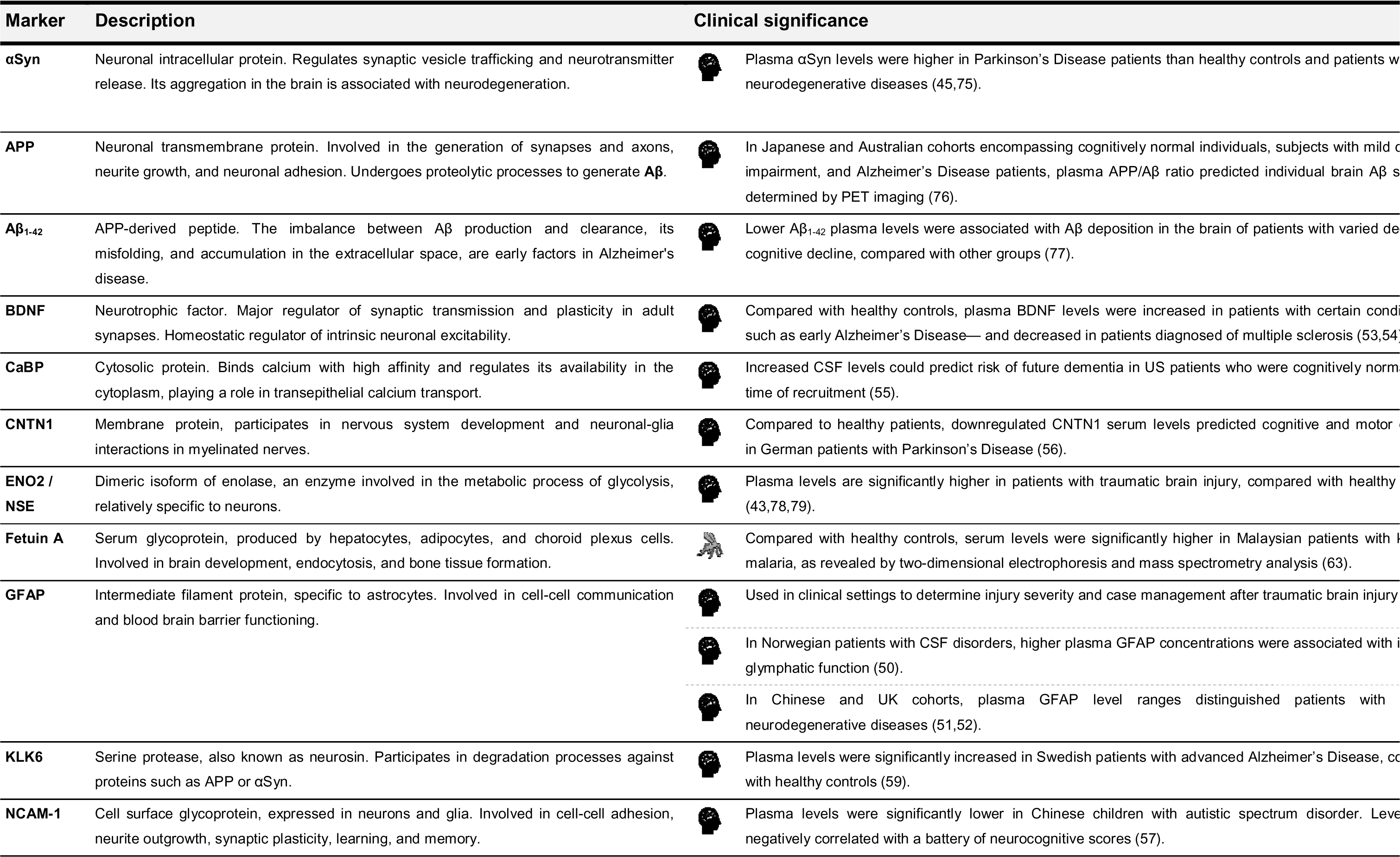
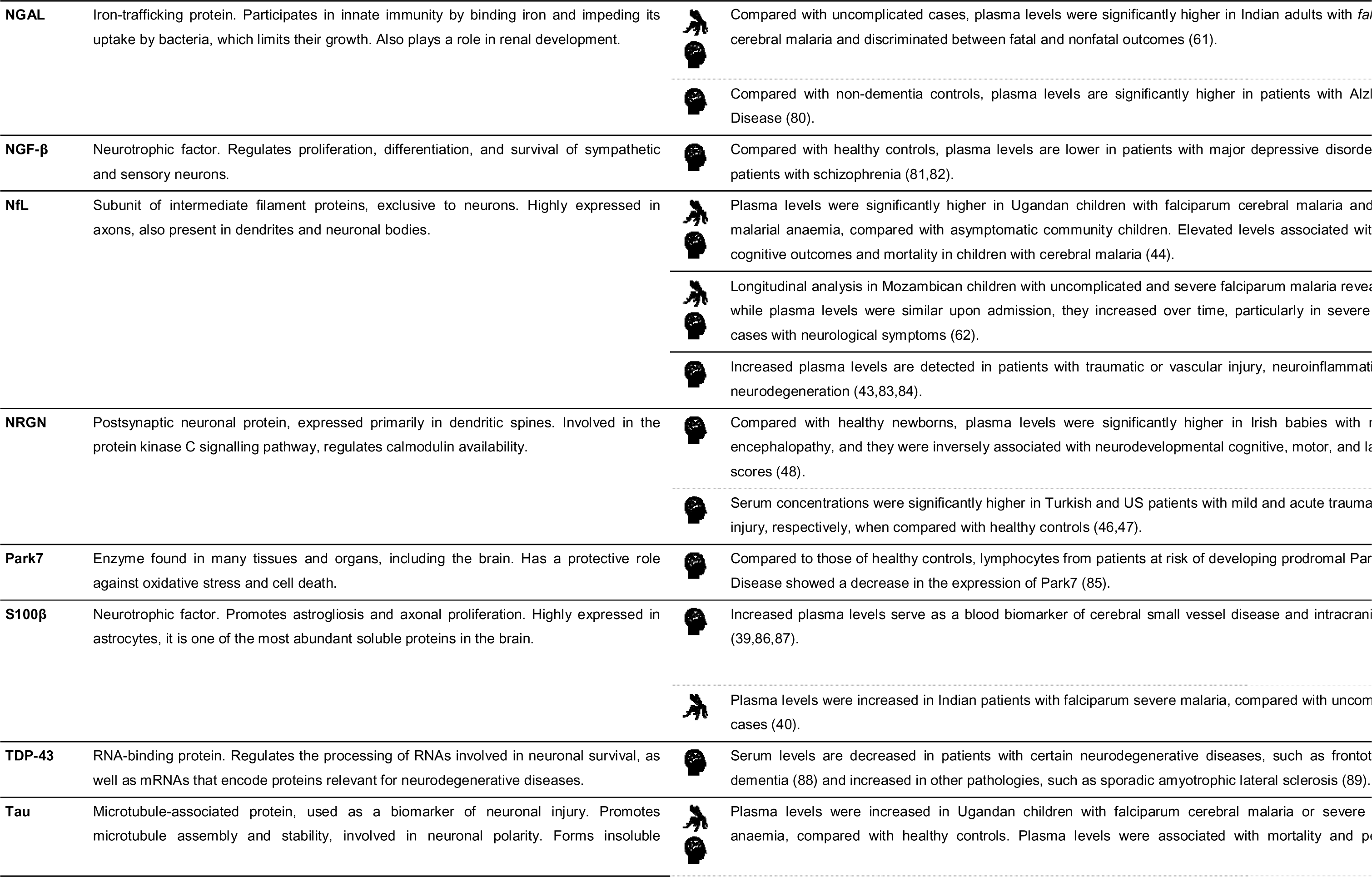
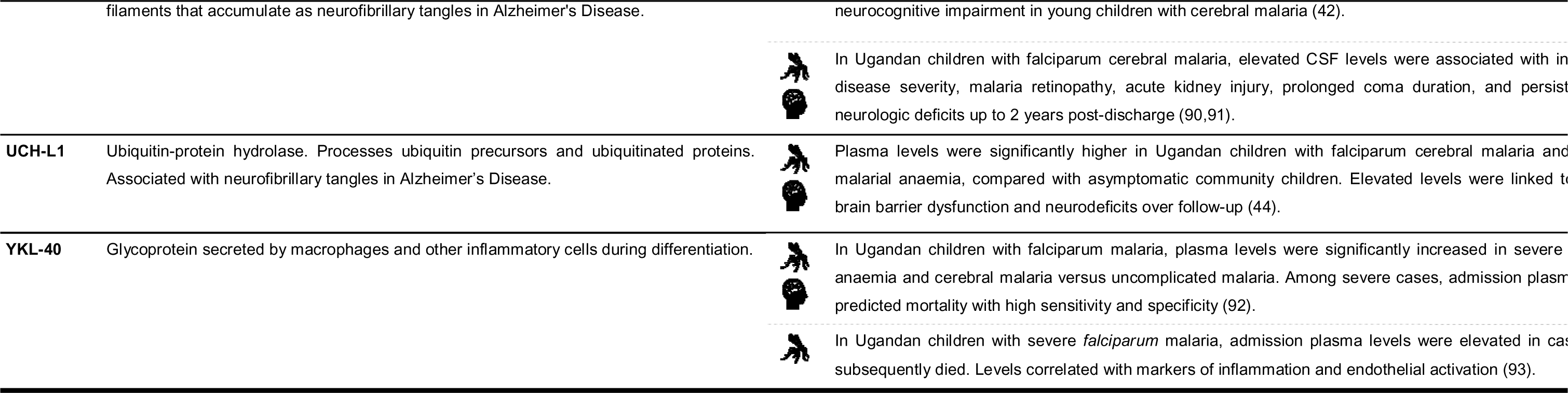
<u>Plasma biomarkers of brain alterations or cerebral injury.</u>. α**Syn**: alpha-Synuclein; **APP**: amyloid-beta precursor protein; **A**β**_1-42_**: amyloid beta (1–42); **BDNF**: brain-derived neurotrophic factor; **CaBP**: Calbindin D; **CNS**: central nervous system; **CNTF**: ciliary neurotrophic factor; **CNTN1**: contactin-1; **CSF**: cerebrospinal fluid; **ENO2** / **NSE**: Enolase 2 / Neuron-specific Enolase; **AHSG**: alpha 2-HS glycoprotein; **FGF-21**: fibroblast growth factor 21; **GDNF**: glial cell line-derived neurotrophic factor; **GFAP**: glial fibrillary acidic protein; **KLK6**: kallikrein 6 / neurosin; **NCAM-1**: neural cell adhesion molecule; **NGAL**: neutrophil gelatinase-associated lipocalin (also known as Lipocalin-2); **NGF-**β: nerve growth factor beta; **NfL**: neurofilament light chain; **NRGN**: neurogranin; **Park7**: Parkinsonism-associated deglycase; **RNA**: ribonucleic acid; **S100**β: S100 calcium-binding protein β; **TDP-43**: TAR DNA-binding protein 43; **Tau**: total Tau protein; **Tau pT181**: phosphorylated Tau protein; **UCH-L1**: ubiquitin carboxy-terminal hydrolase L1; **YKL40**: Chitinase-3-like protein 1. Biomarker descriptions are extracted from **UniProt** (www.uniprot.org, last accessed January 2024). **Legend**: Clinical findings related to: *Plasmodium falciparum* malaria infection; *Plasmodium knowlesi* malaria infection; Neurological conditions.

### 2.3 Measurement of plasma biomarker levels via Luminex and SIMOA assays

Thirty-eight biomarkers were analysed using the customizable Human Luminex® Discovery Assay provided by R&D Biotechne (catalogue ref.: LXSAHM): Alpha-Synuclein (αSyn), Amyloid-beta precursor protein (APP), Angiopoietin-1 (Ang-1), Angiopoietin-2 (Ang-2), Bone Morphogenetic Protein 9 (BMP-9), Calbindin D (CaBP), Chemokine Ligand 2 (CCL2), Chemokine Ligand 4 (CCL4), Chemokine Ligand 5 (CCL5), Chemokine Ligand 18 (CCL18), Contactin-1 (CNTN1), C-Reactive Protein (CRP), Enolase 2 / Neuron-specific Enolase (ENO2/NSE), Fetuin A, Granulocyte-Macrophage Colony-Stimulating Factor (GM-CSF), Interferon Gamma (IFN-γ), Intercellular Adhesion Molecule 1 (ICAM-1), Interleukin 1 alpha (IL-1 α), Interleukin 1 beta (IL-1 β), Interleukin 1RA (IL-1RA), Interleukin 2 (IL-2), Interleukin 4 (IL-4), Interleukin 6 (IL-6), Interleukin 8 (IL-8), Interleukin 10 (IL-10), Interleukin 17 (IL-17A), Lipocalin-2 (NGAL), Myeloperoxidase (MPO), Osteopontin (OPN), Parkinsonism associated deglycase (Park7), Platelet-Derived Growth Factor AA (PDGF-AA) and BB (PDGF-BB), Receptor for Advanced Glycation Endproducts (RAGE), Serine Proteinase Inhibitor!Zclade E1 (Serpin E1), Tumour Necrosis Factor alpha (TNF-α), Vascular Cell Adhesion Molecule (VCAM-1), Vascular Endothelial Growth Factor (VEGF), and von Willebrand Factor A2 domain (vWF-A2).

Additionally, 10 biomarkers were analysed using the Neuroscience 18-plex Human ProcartaPlex™ Panel assay provided by ThermoFisher Scientific Invitrogen™ (catalogue ref.: EPX180-15837-901): Amyloid beta (1–42) (Aβ(_1-42_)), Brain-Derived Neurotrophic Factor (BDNF), Kallikrein 6 (KLK6), Migration Inhibitory Factor (MIF), Nerve Growth Factor beta (NGF-β), Neural Cell Adhesion Molecule (NCAM-1), Neurogranin (NRGN), S100 calcium-binding protein β (S100β), TAR DNA-binding protein 43 (TDP-43), and Chitinase-3-like protein 1 (YKL-40).

Serum levels of biomarkers screened via the Human Luminex® Discovery Assay (#LXSAHM) and the Neuroscience 18-plex Human ProcartaPlex™ Panel (#EPX180-15837-901) were measured using a MAGPIX© bioanalyzer, according to manufacturer’s instructions. Singlicate measurements were taken of each sample. Sample concentrations were extrapolated from a standard curve, fitted using a 6-parameter logistic algorithm. Luminex technology is a multiplex assay platform that allows for the simultaneous measurement of multiple biomarkers in a single plasma sample and can detect molecule concentrations in the picogram range. The use of Luminex technology in the study of biomarkers of brain injury is widely extended (30) due to its high sensitivity and reproducibility.

Lastly, 4 brain injury biomarkers were analysed using the highly sensitive single molecule immunoassay (SIMOA), using the Human Neurology 4-Plex A assay provided by Quanterix (catalogue ref.: 102153): Neurofilament light (NfL), total Tau protein (Tau), Glial Fibrillary Acidic Protein (GFAP), and Ubiquitin Carboxyl-terminal Hydrolase L1 (UCH-L1). Levels of biomarkers screened via the SIMOA Human Neurology 4-Plex A assay (#102153) were measured using a SIMOA HD-X™ Analyzer (Quanterix), according to manufacturer’s instructions. Briefly, samples were thawed at 21°C, and centrifuged at 10,000 RCF for five minutes at the same temperature. On-board the instrument, samples were diluted 1:4 with sample diluent and bound to paramagnetic beads coated with a capture antibody specific for human GFAP, NfL, Tau, and UCH-L1. GFAP, NfL, Tau, and UCH-L1 bound beads were then incubated with a biotinylated GFAP, NfL, Tau, and UCH-L1 detection antibodies in turn conjugated to streptavidin-β-galactosidase complex that acts as a fluorescent tag. Subsequent hydrolysis reaction with a resorufin β-D-galactopyranoside substrate produces a fluorescent signal proportional to the concentration of GFAP, NfL, Tau, and UCH-L1 present. Singlicate measurements were taken of each sample. Sample concentrations were extrapolated from a standard curve, fitted using a 4-parameter logistic algorithm. SIMOA is an ultrasensitive digital immunoassay that allows for the detection and quantification of low-abundance plasma biomarkers with sub-femtomolar sensitivity, high specificity, and high reproducibility (31).

### 2.4 Serological assessment of malaria exposure via Luminex immunoassay

As past infections with *Plasmodium* spp. could also influence observed biomarker levels across participants, total IgG antibody responses of all *Pk*-infected patients and healthy controls were measured using a multiplex bead-based immunoassay built on Luminex® xMAP® technology to assess previous exposure to *P. knowlesi* as well as *P. falciparum, P. vivax, P. malariae*, and *P. ovale*. Sera were screened against a previously optimised panel of 14 blood-stage antigens representing varied markers of malaria exposure, listed in **Supplementary Table 3**. This incorporated 6 *Pk* antigens: *Pk*AMA1 and *Pk*MSP1_19_, historical (long-term) exposure markers that can persist in blood for several years with repeated infections, PkSera3 Ag2 and PkSSP2/TRAP, which have previously been utilised as markers in seroprevalence studies on Malaysian populations (32–34), and *Pk*1 and *Pk*8, exploratory antigens that demonstrated high immunogenicity in preliminary data from assay screenings of pooled sera from *Pk-*infected Malaysian hyperimmune individuals (**K. Tetteh**, **personal communication**). For *P. falciparum* and *P. vivax*, markers of historical exposure, *Pf* / *Pv*MSP1_19_ and *Pf* / *Pv*AMA1, and markers of recent (short-term) exposure known to persist in blood for up to 6-12 months following infection, *Pf*Etramp5 Ag1 and *Pv*RBP 2b, were included (35,36). *Pm*MSP1_19_ and *Po*MSP1_19_ were used to assess historical exposure to *P. malariae* and *P. ovale*, respectively. Additionally, tetanus toxoid vaccine protein from *Clostridium tetani* and glutathione-S-transferase (GST) from *Schistosoma japonicum* were included as non-malaria internal assay controls (37).

The Luminex assay was performed as previously described (38). Briefly, based on previously identified antigen-specific optimal EC_50_ concentrations, each antigen was covalently coupled to a colour-coded MagPlex® bead region (MagPlex, Luminex Corp., Austin, TX) via N-Hydroxysuccinimide/1-Ethyl-3-(3-dimethylaminopropyl)carbodiimide (NHS/EDC) chemistry. Test sera were prepared at 1/400 in diluent buffer (1x Phosphate Buffer Saline (PBS); 0.05% Tween® 20; 0.5% bovine serum albumin; 0.02% sodium azide; 0.1% casein; 0.5% polyvinyl alcohol; 0.5% polyvinylpyrrolidone; 15.25ug/mL *E. coli* lysate) and incubated at 4°C overnight. Antigen-coupled beads were incubated with 50μL of diluted samples and incubated for 90 minutes at room temperature with shaking at 600 RPM before incubation with 50μL of 1/200 R-Phycoerythrin-conjugated AffiniPure F(ab’)₂ Fragment Goat Anti-Human IgG secondary antibody (JacksonImmunoResearch; 109-116-098) for 90 minutes under the same conditions (38).

Pooled sera from hyperimmune individuals in Malaysia (*Pk+*), Tanzania (CP3), and Peru (S1) were included as *Pk, P. falciparum, and P. vivax* positive controls, respectively, in 6-point 5-fold serial dilution curves (1/10 - 1/31250). Commercial WHO reference reagents for anti-*P. falciparum* (10/198) human serum and anti-*P. vivax* (19/198) human plasma were also added as positive controls at 1/400 and 1/4000. Public Health England (PHE) malaria naïve human sera (n=30) were included as negative controls at 1/400. Two wells of diluent buffer served as blank controls to allow subtraction of background signal. The plates were read using the MAGPIX© instrument (Luminex Corp., Austin, TX) with the raw data recorded as Median Fluorescent Intensity (MFI) values using an acquisition target of ≥30 beads per region per well. The data were background-adjusted and normalised as described by Wu *et al* (38).

### 2.5 Statistical analysis

Sociodemographic characteristics of the participants were reported as mean (±SD) for continuous data, or absolute frequencies (%) for categorical data. Continuous data was assessed for normality using Shapiro-Wilk tests. Comparisons between *Pk*-infected cases and healthy controls were made using *t*-tests or Wilcoxon tests for normally and non-normally distributed continuous data, respectively, and χ^2^ tests for categorical data.

Differences in levels of individual biomarkers between *Pk*-infected patients and controls were examined using two-tailed *t*-tests and Wilcoxon tests for normally and non-normally distributed data, respectively. Bonferroni correction for multiple comparisons was applied. Correlation matrixes were used to visualize the relationships between biomarker levels in both groups, with previous data scaling. Any potential correlations between biomarker levels and demographic and clinical parameters such as age and parasitemia were evaluated using Spearman’s rank correlation coefficient.

To elucidate potential profiles of biomarker levels that could distinguish between infected patients and controls, a hierarchical clustering heatmap analysis was performed, prior scaling of biomarker data. This provided a visual representation of the relationships between individuals based on the similarity of their biomarker profiles. Initially, the analysis was conducted on the entire set of biomarkers to comprehensively explore the clustering patterns within the dataset. Subsequently, the clustering analysis was applied separately to subsets of biomarkers representing brain injury, infection and immunity, and vascular biomarkers.

To assess malaria exposure, a hierarchical clustering heatmap was similarly performed to allow visual comparison of individual antibody responses to *Pk-*specific antigen profiles across study participants. Comparisons of group means of MFI responses between *Pk*-infected cases and healthy controls were conducted using the non-parametric Kruskal-Wallis test with Dunn’s correction for multiple comparisons applied.

Statistical analyses were performed in R (version 4.3.1) with RStudio software (version 2023.09.1-494). Data was visualized in R (version 4.3.1) with RStudio software (version 2023.09.1-494) and GraphPad Prism 10. P values of less than 0.05 (two-tailed) were considered statistically significant.

## 3 Results

### 3.1 Cohort characteristics

All participants in our sub-study were male (N=38, 100%). Both *Pk*-infected patients and community-matched, age-matched uninfected controls had an average age of 39 (±15) years. In the *Pk*-infected group, patients presented with median parasitemia 15,200.00 parasites/μL and 26,746.00 interquartile range.

### 3.2 Concentrations of individual markers between *Pk*-infected patients and controls

#### Biomarkers of brain injury

Bonferroni-corrected analyses revealed significantly higher plasma levels of the following brain injury biomarkers in the *Pk*-infected group, compared with uninfected controls: Tau (p=0.0007), UCH-L1 (p<0.0001), αSyn (p<0.0001), Park7 (p=0.0006), NRGN (p=0.0022), and TDP-43 (p=0.005). In contrast, levels of the following biomarkers were found to be significantly lower in the *Pk*-infected group when compared with uninfected controls: CaBD (p<0.0001), CNTN1 (p<0.0001), NCAM-1 (p<0.0001), BDNF (p<0.0001), GFAP (p=0.0013), and KLK6 (p=0.0126). Results obtained on CaBD and CNTN1 levels revealed clear cutoff values that allowed for subject classification based on infection status: All *Pk*-infected patients (19/19, 100%) presented with plasma CaBD levels below 1,400pg/mL or CNTN1 levels below 12,500pg/mL, whereas concentrations in all uninfected controls (19/19, 100%) were above 1,400pg/mL for CNTN1 and above 15,000pg/mL for CNTN1, respectively (**Figure 2**, **Supplementary Table 4**).

**Figure 2.**
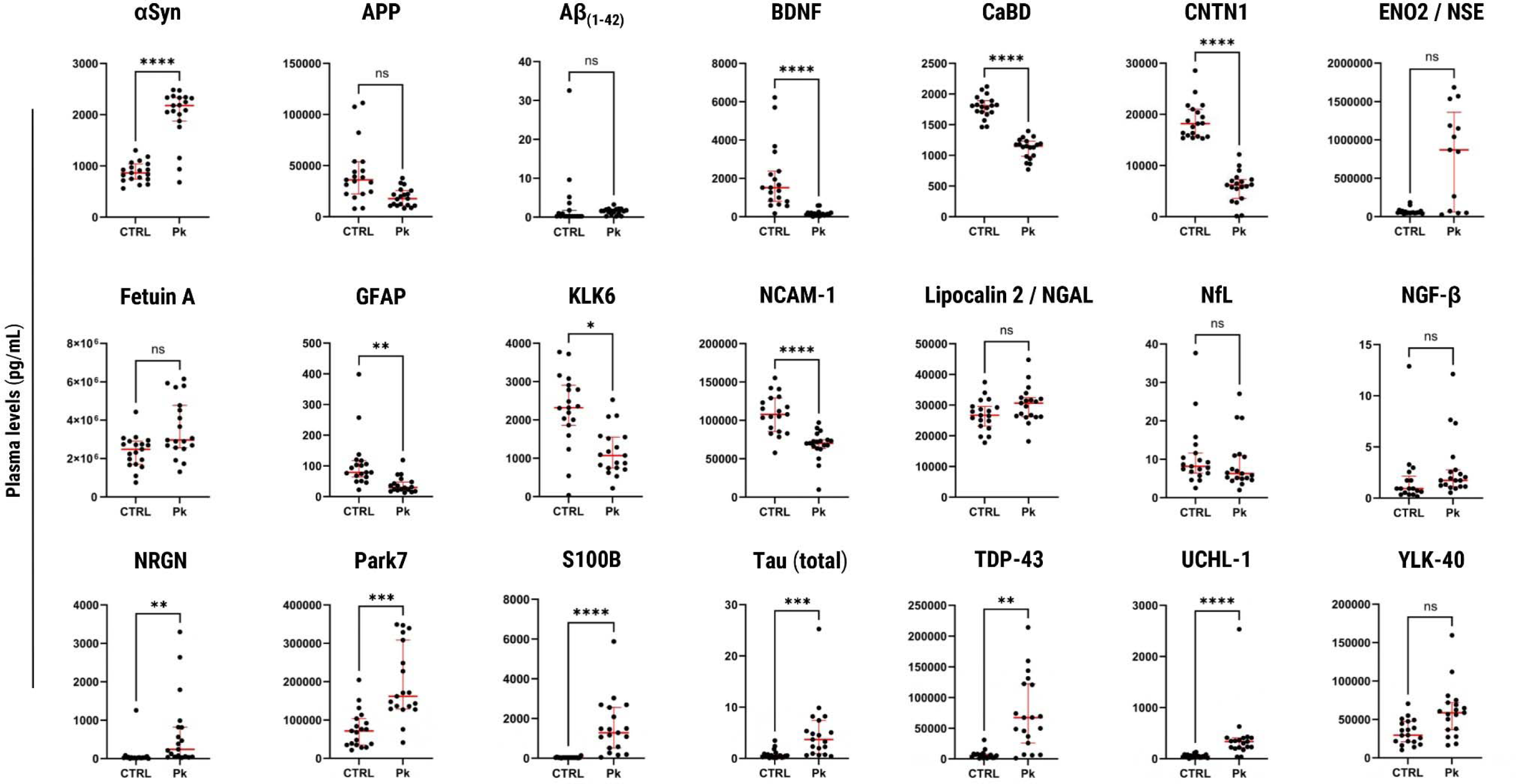
<u>Group comparisons: Levels of blood circulating biomarkers of brain injury.</u>. α**Syn**: alpha-Synuclein; **APP**: amyloid-beta precursor protein; **A**β**_1-42_**: amyloid beta (1–42); **BDNF**: brain-derived neurotrophic factor; **CaBP**: Calbindin D; **CNS**: central nervous system; **CNTF**: ciliary neurotrophic factor; **CNTN1**: contactin-1; **CSF**: cerebrospinal fluid; **ENO2** / **NSE**: Enolase 2 / Neuron-specific Enolase; **AHSG**: alpha 2-HS glycoprotein; **FGF-21**: fibroblast growth factor 21; **GDNF**: glial cell line-derived neurotrophic factor; **GFAP**: glial fibrillary acidic protein; **KLK6**: kallikrein 6 / neurosin; **NCAM-1**: neural cell adhesion molecule; **NGAL**: neutrophil gelatinase-associated lipocalin (also known as Lipocalin-2); **NGF-**β: nerve growth factor beta; **NfL**: neurofilament light chain; **NRGN**: neurogranin; **Park7**: Parkinsonism-associated deglycase; **RNA**: ribonucleic acid; **S100**β: S100 calcium-binding protein β; **TDP-43**: TAR DNA-binding protein 43; **Tau**: total Tau protein; **Tau pT181**: phosphorylated Tau protein; **UCH-L1**: ubiquitin carboxy-terminal hydrolase L1; **YKL40**: Chitinase-3-like protein 1. Wilcoxon test results: *p<0.05; **p<0.01; ***p<0.001; ****p<0.0001; ns: no significance.

Since S100B and Aβ(_1-42_) plasma levels were below the range of detection in most uninfected subjects, we first compared the proportion of individuals with detectable levels of these biomarkers in each group using a Chi-square test with Yates’ correction. The percentage of participants with plasma levels within the range of detection was significantly higher in the *Pk*-infected group (19/19, 100%) than in the uninfected control group (4/19, 21.05%) (p<0.0001). Similarly for Aβ(_1-42_), the proportion of individuals with detectable plasma levels in the *Pk*-infected group (17/19, 89.47%) was significantly higher compared with their uninfected peers (7/19, 36.84%) (p= 0.0025).

To enable group comparisons for these two biomarkers, participants with values below the range of detection were assigned a numerical value corresponding to half the value of the lower threshold of detection. According to the manufacturer’s information, these values were 27.31 pg/mL for S100B and 0.22 pg/mL for Aβ(_1-42_). Achieved this, Wilcoxon tests revealed significantly higher plasma S100B levels in the *Pk*-infected group, which survived Bonferroni correction for multiple comparisons (p<0.0001), whereas no significant group differences were found for Aβ(_1-42_) plasma levels.

#### Biomarkers of infection and immune activation

Group differences in plasma levels were found after correction for multiple comparisons. IL-1RA (p<0.0001), IL-10 (p<0.0001), and MPO levels (p=0.0314) were significantly higher in the *Pk*-infected group when compared with their uninfected peers, whereas CCL4 (p<0.0001), CCL5 (p<0.0001), CRP (p=0.0280), and RAGE levels (p=0.0025) were significantly lower (**Supplementary Figure 1**, **Supplementary Table 4**).

#### Biomarkers of vascular activation and injury

Ang-2/Ang-1 ratios (p<0.0001) and VCAM-1 levels (p=0.0001) were significantly higher in *Pk*-infected patients compared with uninfected controls, while Ang-1 (p<0.0001), BMP-9 (p<0.0001), PDGF-AA and -BB (p<0.0001), and Serpin E1 levels (p<0.0001) were significantly lower (**Supplementary Figure 2**, **Supplementary Table 4**).

### 3.3 Hierarchical clustering of samples based on biomarker blood levels

Hierarchical clustering heatmap analyses revealed distinct group profiles for brain injury biomarkers (**Figure 3A**). Most infected individuals clustered together (17/19, 89.47%), indicating a cohesive pattern of elevated levels of Park7, S100B, αSyn, and TDP-43. Two infected individuals exhibited atypical clustering with the healthy controls, suggesting a subgroup with a distinct biomarker profile. In the control group (19/19, 100%), certain biomarkers, including BDNF, CaBD, CNTN1, and GFAP, displayed higher levels and clustered together, representing a baseline biomarker profile in healthy individuals. As mentioned, two infected individuals (2/19, 10.53%) clustered with the control group in this category, indicating potential overlap or similarity in the levels of these specific biomarkers between infected and control individuals.

**Figure 3.**
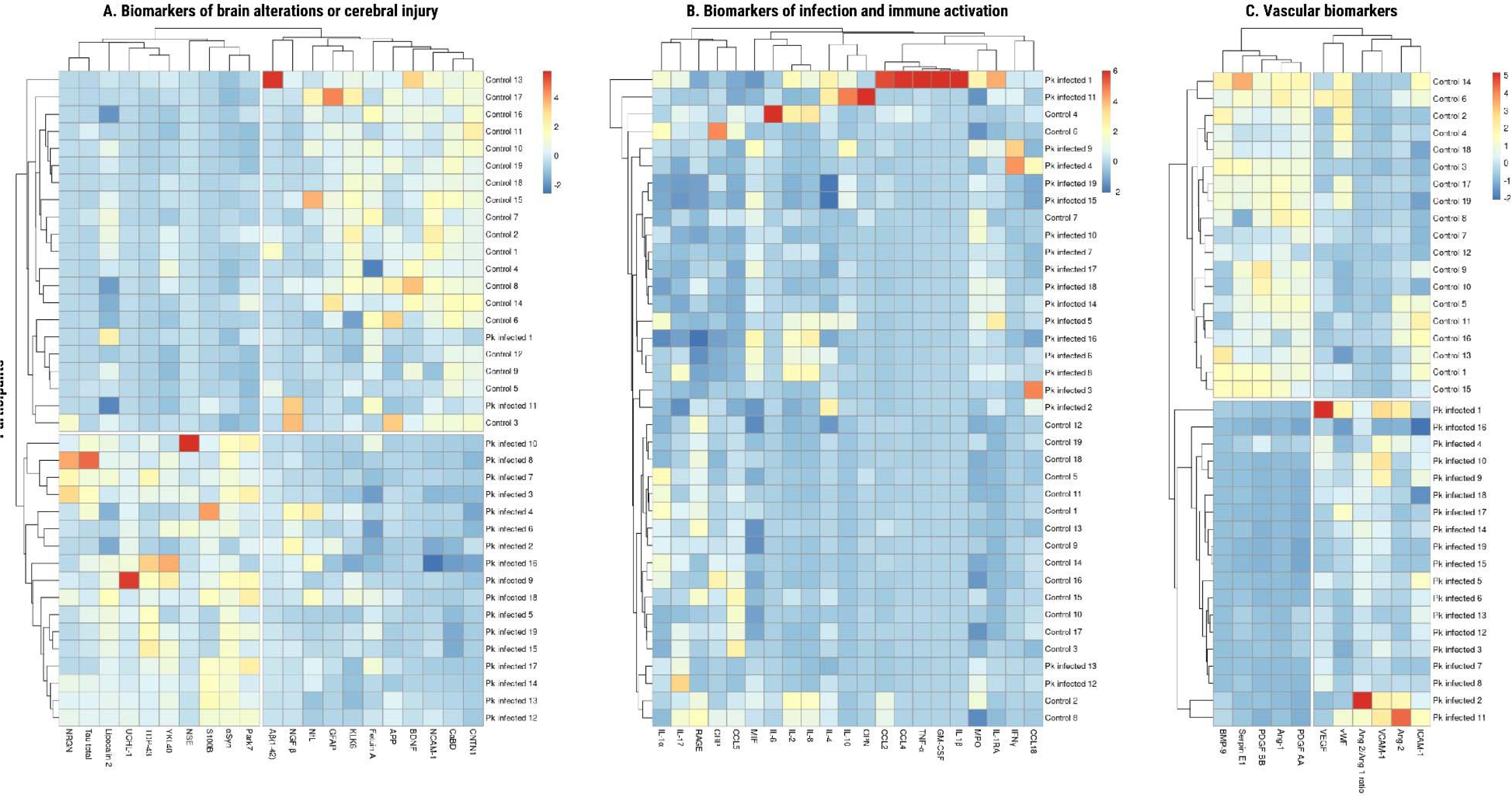
<u>Hierarchical clustering heatmaps: Distinct circulating biomarker profiles between groups.</u>. **Fig 3A. Biomarkers of brain alterations or cerebral injury**: α**Syn**: alpha-Synuclein; **APP**: amyloid-beta precursor protein; **A**β**_1-42_**: amyloid beta (1–42); **BDNF**: brain-derived neurotrophic factor; **CaBP**: Calbindin D; **CNTN1**: contactin-1; **CSF**: cerebrospinal fluid; **ENO2** / **NSE**: Enolase 2 / Neuron-specific Enolase; **GFAP**: glial fibrillary acidic protein; **KLK6**: kallikrein 6 / neurosin; **NCAM-1**: neural cell adhesion molecule; **Lipocalin-2**: neutrophil gelatinase-associated lipocalin; **NGF-**β: nerve growth factor beta; **NfL**: neurofilament light chain; **NRGN**: neurogranin; **Park7**: Parkinsonism-associated deglycase; **S100B**: S100 calcium-binding protein β; **TDP-43**: TAR DNA-binding protein 43; **Tau**: total Tau protein; **UCH-L1**: ubiquitin carboxy-terminal hydrolase L1; **YKL40**: Chitinase-3-like protein 1. **Fig 3B. Biomarkers of infection and immune activation**: **CCL2**: chemokine (C-C motif) ligand 2; **CCL4**: chemokine (C-C motif) ligands 4; **CCL5**: chemokine (C-C motif) ligand 5; **CCL18**: chemokine (C-C motif) ligand 18; **CRP**: C-reactive protein; **GM-CSF**: granulocyte-macrophage colony-stimulating factor; **IFN-**γ: interferon gamma; **IL-1**α: interleukin 1 alpha; **IL-1**β: interleukin 1 beta; **IL-1RA**: interleukin 1RA; **IL-2**: interleukin 2; **IL-4**: interleukin 4; **IL-6**: interleukin 6; **IL-8**: interleukin 8; **IL-10**: interleukin 10; **IL-17A**: interleukin 17; **MIF**: migration inhibitory factor; **MPO**: myeloperoxidase; **OPN**: osteopontin; **RAGE**: receptor for advanced glycation end-products; **TNF-**α: tumour Necrosis Factor alpha. **Fig 3C. Vascular biomarkers**: **Ang-1**: angiopoietin-1; **Ang-2**: angiopoietin-2; **Ang-2/Ang-1**: ratio between Ang-2 and Ang-1; **BMP-9**: bone morphogenetic protein 9; **ICAM-1**: intercellular adhesion molecule 1; **PDGF-AA**: platelet-derived growth factor AA; **PDGF-BB**: platelet-derived growth factor BB; **Serpin E1**: Serine Proteinase Inhibitorl71clade E1; **VCAM-1**: vascular cell adhesion molecule; **VEGF**: vascular endothelial growth factor; **vWF-A2**: von Willebrand Factor (A2 domain). Data is scaled.

Clustering of biomarkers associated with infection and immune activation did not reveal distinct separation between the two groups (**Figure 3B**). Higher levels of IL-1RA and MPO predominantly clustered in the infected group (17/19, 89.47%), and two control individuals also exhibited this pattern (2/19, 10.53%). One infected individual (1/19, 5.26%) displayed very high levels of IL-1β, GM-CSF, TNF-α, CCL4, and CCL2. Another infected individual (1/19, 5.26%) exhibited very high levels of OPN and IL-10. Additionally, one control individual showed high levels of IL-6 (1/19, 5.26%).

Lastly, clustering of vascular biomarkers revealed clear and distinctive group profiles (**Figure 3C**). In the control group (19/19, 100%), a cohesive cluster formed by high levels of PDGF AA, Ang-1, PDGF BB, Serpin E1, and BMP-9 was observed. Conversely, in the infected group (19/19, 100%), a distinct cluster of higher levels of VCAM-1 and the Ang-2/Ang-1 ratio was identified.

Correlations between biomarkers in each group can be observed in **Supplementary Figure 3**.

### 3.4 Correlation of biomarker levels with age and parasitemia

No significant correlations were identified between the levels of any of the examined biomarkers and the age of participants within each respective group, as illustrated in **Supplementary Figure 4**. Furthermore, within the *Pk*-infected group, no significant correlations were observed between biomarker levels and the percentage of parasitemia (**Supplementary Figure 5**).

### 3.5 Serological markers of previous malaria exposure

Overall, the majority of participants generated low antibody responses across *Pk, P. vivax, P. falciparum, P. ovale*, and *P. malariae* antigens with the exception of a few high responders (**Supplementary Figure 6**). Clustering of exposure markers in the hierarchical heatmap analysis did not reveal distinct separation between responses of *Pk*-infected patients and uninfected controls (**Supplementary Figure 7**). Among *Pk* antigens, uninfected controls exhibited higher responses against *Pk*8 (p<0.0001), *Pk*SERA3 Ag2 (p=0.0406), and *Pk*1 (p=0.0250) than *Pk*-infected patients, but no differences were observed for *Pk*AMA1 (p>0.9999), *Pk*MSP1_19_ (p=0.7778), or *Pk*SSP2 (p=0.2592).

## 4 Discussion

Our study sheds light on the potential neurological implications of *Pk* infection in Malaysia. Through the investigation of circulating levels of brain injury, inflammation, and vascular biomarkers in *Pk*-infected patients compared to healthy controls, we uncovered compelling evidence suggesting a significant impact of *Pk* infection on brain health. In line with previous reports suggesting neurological involvement in severe malaria, our findings demonstrate increased levels of several biomarkers associated with cerebral injury in *Pk*-infected patients. Notably, elevated levels of S100B, Tau, UCH-L1, αSyn, Park7, NRGN, and TDP-43 signify potential brain damage in response to *Pk* infection in our cohort.

Alterations in circulating levels of S100B, Tau, and UCH-L1 have been documented in previous investigations of severe malaria. S100B, an abundant neurotrophic factor predominantly expressed in astrocytes, serves as a biomarker of blood-brain barrier permeability and central nervous system injury (39). Our group reported increased plasma levels of S100B in Indian patients with falciparum severe malaria compared to uncomplicated cases (40). Similarly, levels of microtubule-associated protein Tau, a neuropathological hallmark of Alzheimer’s Disease and a biomarker for brain injury (41), were found increased in the plasma of Ugandan children with severe falciparum malarial anemia or cerebral malaria compared to uninfected controls. In cerebral malaria cases, Tau levels correlated with mortality and persistent neurocognitive impairment in survivors (42). Lastly, plasma levels of the ubiquitin-protein hydrolase UCH-L1, a marker of neuronal damage (43) were significantly higher in Ugandan children with falciparum cerebral malaria and severe malarial anemia compared with asymptomatic community children. The elevated plasma levels were associated with surrogate markers of blood-brain barrier dysfunction and cognitive deficits over follow-up (44). Our findings of increased S100B, Tau, and UCH-L1 plasma levels during *Pk* infection align with these observations, suggesting potential cerebral involvement, neuronal injury, and blood-brain barrier disruption in knowlesi malaria pathogenesis.

In broader clinical contexts, elevated circulating biomarkers in our *Pk*-infected cohort have been associated with diverse conditions and are used as indicators of brain injury or dysfunction. For instance, patients with Parkinson’s Disease had significantly higher plasma levels of synaptic vesicle trafficking regulator αSyn compared to healthy controls (45), and elevated levels of post-synaptic protein NRGN were associated with mild to acute traumatic brain injury (46,47), as well as neonatal encephalopathy (48). The increased levels of αSyn and NRGN in our *Pk*-infected patients suggest potential neurological implications, prompting further investigation into the underlying mechanisms of knowlesi malaria pathogenesis and their clinical significance over time.

In our study, several biomarkers were significantly decreased in *Pk*-infected individuals compared to healthy controls, including GFAP, BDNF, CaBD, CNTN1, NCAM-1, and KLK6. The astrocytic intermediate filament protein GFAP, used in clinical settings to assess severity of traumatic brain injury (49), glymphatic function (50), and neurodegeneration (51,52), was not found to be elevated in Ugandan children with falciparum cerebral malaria and severe malarial anemia (44). In contrast, the observed differences in GFAP levels in our groups may suggest altered astroglial and/or glymphatic functions in infected participants, supporting our hypothesis of cerebral involvement in *Pk* malaria. Conversely, plasma levels of the neurotrophic factor BDNF, reported to be increased in early Alzheimer’s disease (53) and decreased in multiple sclerosis (54), were also lower in our study participants, potentially indicating dysregulated neuroprotective mechanisms during *Pk* infection. Furthermore, increased cerebrospinal fluid levels of CaBD, a protein involved in calcium homeostasis and neuronal signalling, were associated with risk of dementia in cognitively normal patients (55). CNTN1 and NCAM-1, two crucial factors for neural development and synaptic formation, were decreased in our *Pk*-infected patients. Downregulated CNTN1 serum levels predicted cognitive and motor declines in patients with Parkinson’s Disease (56), while lower plasma NCAM-1 levels correlated with cognitive impairment (57,58). Lastly, plasma levels of the neuroinflammation modulator KLK6 were significantly increased in patients with advanced Alzheimer’s Disease compared with healthy controls (59). Although challenging to interpret, these results suggest cerebral involvement during *Pk* infection and a potential risk of cognitive decline, indicating a complex interplay between neuroinflammatory and neuroprotective mechanisms that warrant further investigation.

Despite our expectations based on existing malaria literature (**Table 1**), we did not observe significant group differences in levels of some biomarkers, such as the iron-trafficking protein Lipocalin-2 (NGAL), a recognized biomarker of neuroinflammation (60). Elevated plasma NGAL levels were associated with cerebral malaria in adult patients from India and distinguished between fatal and nonfatal outcomes (61). Similarly, plasma NfL levels in Mozambican children with uncomplicated and severe falciparum malaria showed no baseline group differences, but a significant increase over time, particularly in severe malaria cases with neurological symptoms, suggesting NfL as a potential follow-up biomarker of brain injury in malaria (62). Additionally, the glycoprotein Fetuin A was previously reported as elevated in serum from Malaysian patients with *Pk* malaria compared to uninfected controls (63). Lastly, plasma levels of the neurotrophic factor CNTF were significantly reduced in Thai patients with cerebral malaria or severe malaria with renal failure, compared to severe malaria cases without cerebral or renal involvement (64). Our findings, diverging from anticipated outcomes, underscore the complexity of biomarker dynamics in infections with different *Plasmodium* species and emphasize the need for further exploration in the context of *Pk* malaria.

In our hierarchical clustering analysis, we observed distinct group profiles for brain injury biomarker levels, indicating differences between *Pk*-infected patients and healthy controls. Most infected individuals clustered together with elevated levels of specific biomarkers such as S100B, αSyn, Park7, and TDP-43, suggesting a cohesive pattern of cerebral injury. However, two infected individuals exhibited an atypical clustering with the healthy controls, hinting at potential subgroup variability within the infected cohort, or different stages of disease progression. Conversely, in the control group, several biomarkers including BDNF, GFAP, CaBD, and CNTN1 displayed higher levels and clustered together, representing a baseline biomarker profile in healthy individuals. As mentioned, two infected individuals clustered with the control group in this category, indicating potential overlap or milder infection.

Clustering of vascular biomarkers revealed clear and distinctive group profiles. Uninfected individuals formed a cohesive cluster with high levels of PDGF AA, Ang-1, PDGF BB, Serpin E1, and BMP-9, revealing a healthy, baseline vascular profile. Conversely, the infected group cluster had high levels of VCAM-1 and Ang-2/Ang-1 ratio, indicative of vascular involvement and endothelial activation during *Pk* infection. The elevated Ang-2/Ang-1 ratio observed in our infected group aligns with findings from previous studies on severe malaria caused by other *Plasmodium* species (65–70). Similarly, increased levels of VCAM-1 have been reported in both falciparum and vivax malaria infections (71,72). In contrast to brain injury and vascular biomarker profiles, hierarchical clustering of biomarkers associated with infection and immune activation did not reveal distinct separation between the two groups.

Lastly, total IgG antibody responses observed against *Plasmodium* spp. antigens, including *Pk*, were low not only among most uninfected controls but also *Pk-*infected patients, and did not form distinct clusters between the two groups. This suggests the cohort was largely malaria naïve, with low reactivity to long-term infection markers *Pk*/*Pv*/*Pf*/*Pm*/*Po*MSP1_19_ and *Pk*/*Pv*/*Pf*AMA1 observed at levels comparable to the PHE malaria naïve controls in all but two *Pk*-infected patients (73). As naturally acquired responses against these antigens develop cumulatively with repeated exposure, this indicates the occurrence of very few historical *Plasmodium* infections across participants (74). This could highlight a lack of clinical immunity, rendering *Pk-*infected patients more susceptible to potential malaria complications reflected in the brain and vascular biomarker profiles in this study, though this was not possible to investigate due to insufficient clinical information. The lack of elevated responses to *Pk* antigens SSP2, SERA3 Ag2, *Pk*1 and *Pk*8 may indicate there was inadequate time for *Pk-*infected patients to mount a response prior to the sampling period (32).

Among uninfected controls, serological analyses identified two individuals exhibiting notably higher responses: one control to *Pk* exploratory marker *Pk*8 and *P. vivax* short-term exposure marker *Pv*RBP2b, and another also to *Pv*RBP2b. Although this suggests previous exposures to *Pk* and *P. vivax*, this did not appear to explain any atypical deviations in plasma biomarker levels observed in a minority of individuals in this group.

Our study had several limitations. Firstly, the lack of detailed clinical information regarding malaria severity and the presence or absence of renal failure among the *Pk*-infected group restricts our ability to fully contextualize the observed alterations in brain injury biomarkers. While recent studies have implicated AKI in the pathogenesis of brain injury (40) and long-term neurocognitive sequelae (10,11), we were unable to directly investigate this relationship. Since AKI is a common complication of knowlesi malaria (27), it is likely that similar distant organ pathways are involved, resulting in an exacerbation of the brain changes (26). Secondly, our study did not include long-term follow-up assessments to determine whether the elevated biomarker levels observed in *Pk*-infected patients return to baseline or exhibit a steady increase over time. This would have been particularly relevant for NfL, which increases in the weeks following cerebral insult and could provide valuable insights into the persistence or resolution of brain injury following *Pk* infection (43,62). Lastly, a symptomatic, non-malaria control group would allow stronger ascertainment of associations between biomarkers and *Pk* infection.

In conclusion, our study represents the first comprehensive exploration of surrogate markers of cerebral involvement in *Pk*-infected patients from Malaysia. Through the investigation of circulating levels of brain injury, inflammation, and vascular biomarkers in *Pk*-infected patients compared to healthy controls, we uncovered compelling evidence suggesting a significant impact of *Pk* infection on brain and vascular health. The identification of distinct patterns in the clustering of biomarkers associated with cerebral injury and endothelial dysfunction highlights the complex interplay between these pathways during *Pk* infection. Our study lays the groundwork for further research to elucidate the pathophysiology of this disease, and its potential long-term effects on the brain. Additional investigations are urgently needed to assess structural and functional changes in the brain during knowlesi malaria, through both neuroimaging and longitudinal neurocognitive evaluations. Our findings not only contribute to our understanding of *Pk* malaria but may also have future implications for the diagnosis, management, and therapeutic targeting of neurological complications associated with this emerging infectious disease.

## 5 Ethics approval and consent

This study has been reviewed and approved by the Medical Research and Ethics Committee of the Ministry of Health in Malaysia (reference NMRR ID 22-02557-1KV) and by the Observational/Interventions Research Ethics Committee at the London School of Hygiene and Tropical Medicine, UK (reference 27902). The shipment of samples from Malaysia to the UK was performed in compliance with national and international regulations in both sites. The samples are registered with the Human Tissue Authority, in accordance with UK national guidelines.

## Data Availability

All data produced in the present work are contained in the manuscript.

## 7 Author Contributions

**CBC**: conceptualization, methodology, investigation, formal analysis and interpretation, visualization, writing – original draft, review & editing. **ED**: methodology, formal analysis and interpretation, visualization, writing – review & editing. **NRN**: conceptualization, methodology, investigation, writing – review & editing. **MYL**: conceptualization, methodology, investigation, writing – review & editing. **KT**: methodology, investigation, resources, and writing – review & editing. **CD**: methodology, investigation, resources, and writing – review & editing. **SK**: conceptualization, investigation, and writing – review & editing. **YLL**: conceptualization, methodology, investigation, resources, supervision, and writing – review & editing. **SCW**: conceptualization, methodology, investigation, resources, supervision, and writing – review & editing. All authors approved the final version.

## 9 Acknowledgments

We extend our gratitude to the local health officers and personnel from Mersing, Kota Tinggi, and Pejabat Kesihatan Dareah Kota Tinggi (Johor), Kuala Lipis and Kesihatan Betau (Pahang), Sungai Siput (Perak), Kuala Kubu Bharu (Selangor), and Setiu (Trengganu) for their invaluable assistance with sampling. We express our sincere appreciation to Dr Christian Bottomley, Ana Chopo-Pizarro, and Helena Brazal Monzó at LSHTM for their invaluable insights, which greatly enhanced the quality of this research.

