## Supplementary figures and images for "*Plasmodium knowlesi* infection is associated with elevated circulating biomarkers of brain injury and endothelial activation"

### Supplementary Figure 1

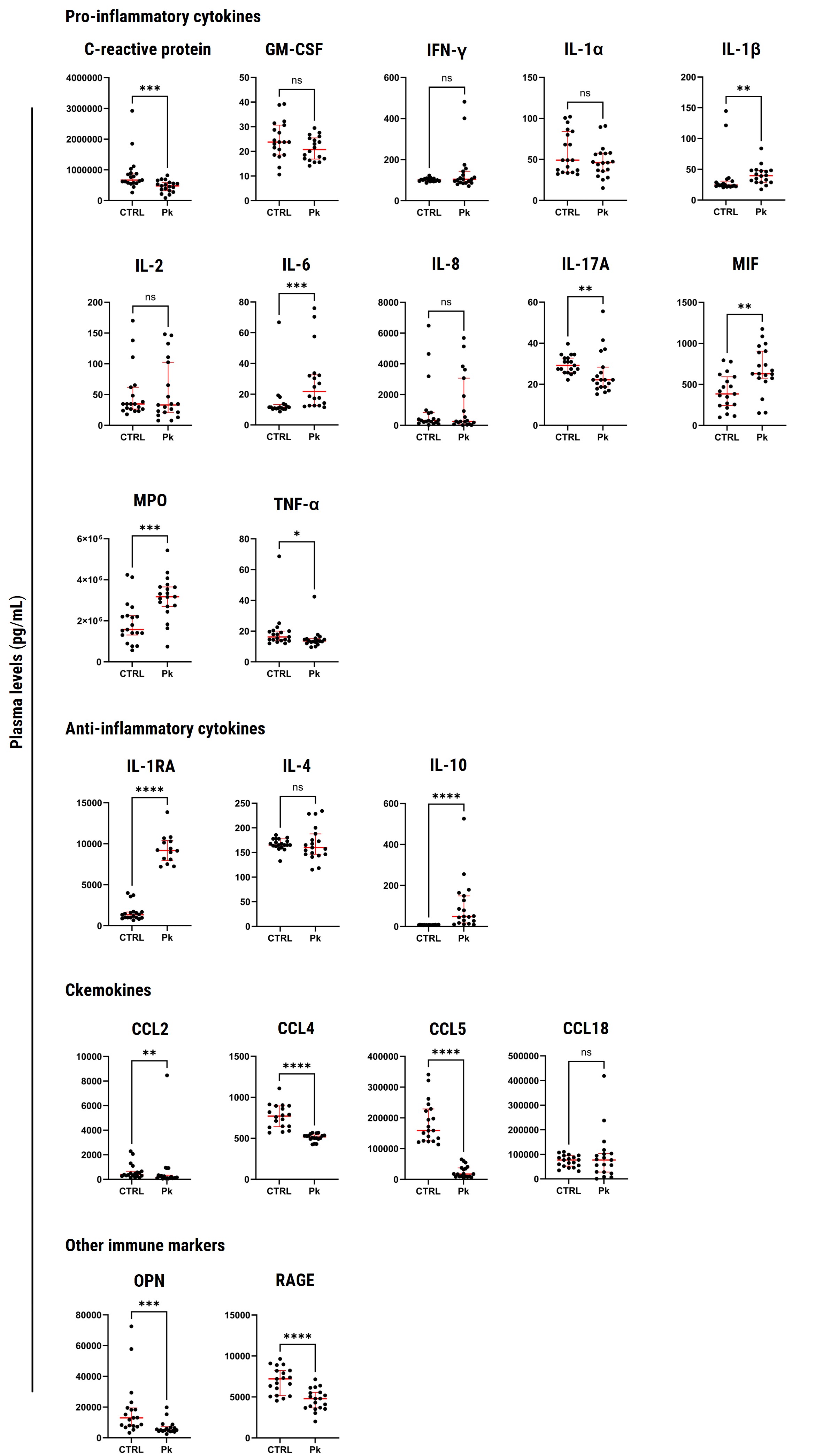

### Supplementary Figure 2

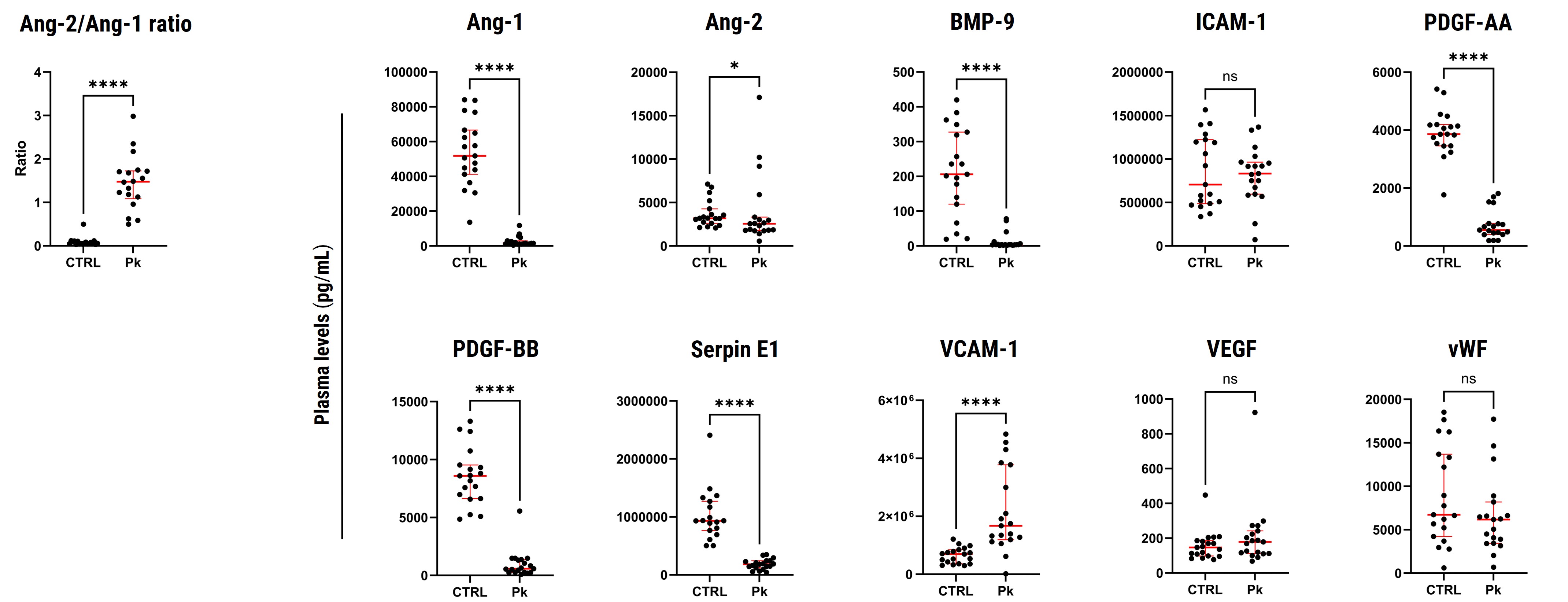

### Supplementary Figure 3

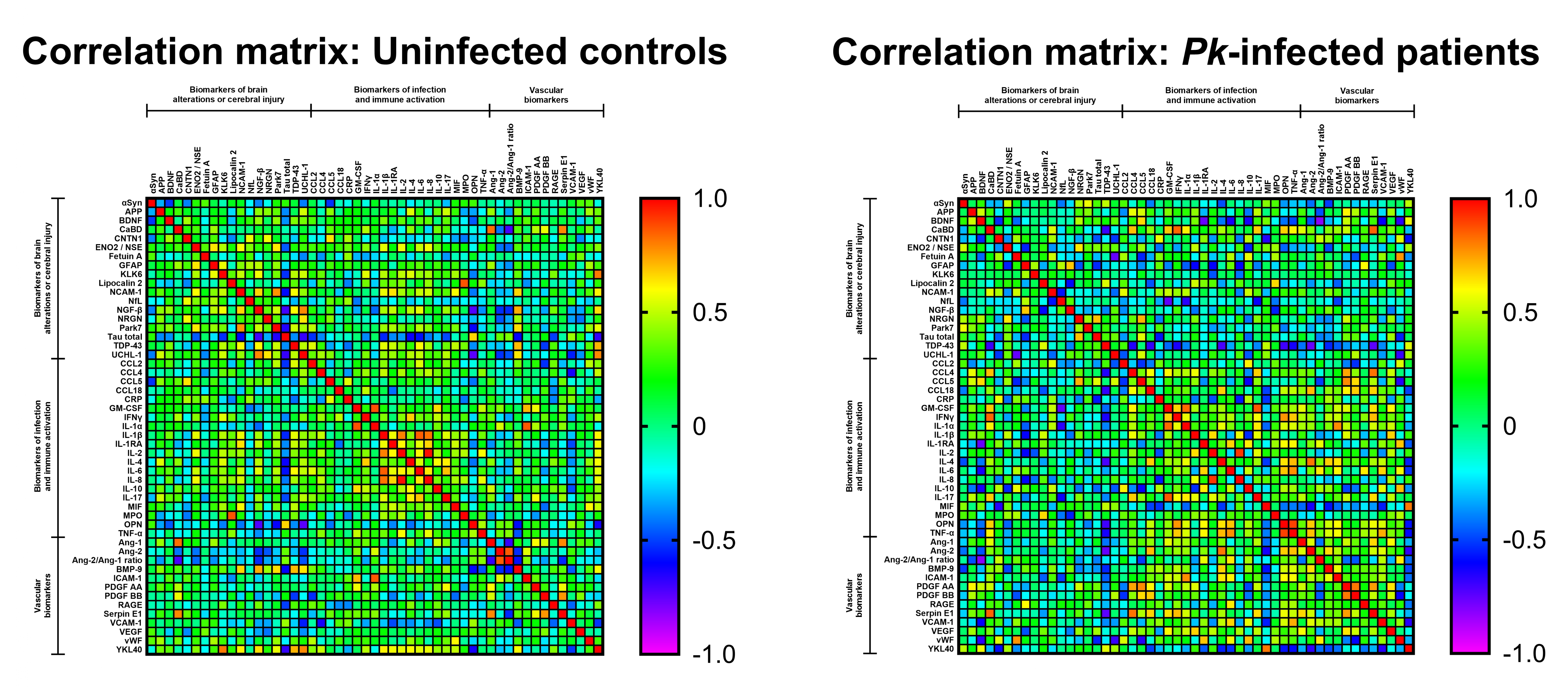

### Supplementary Figure 4

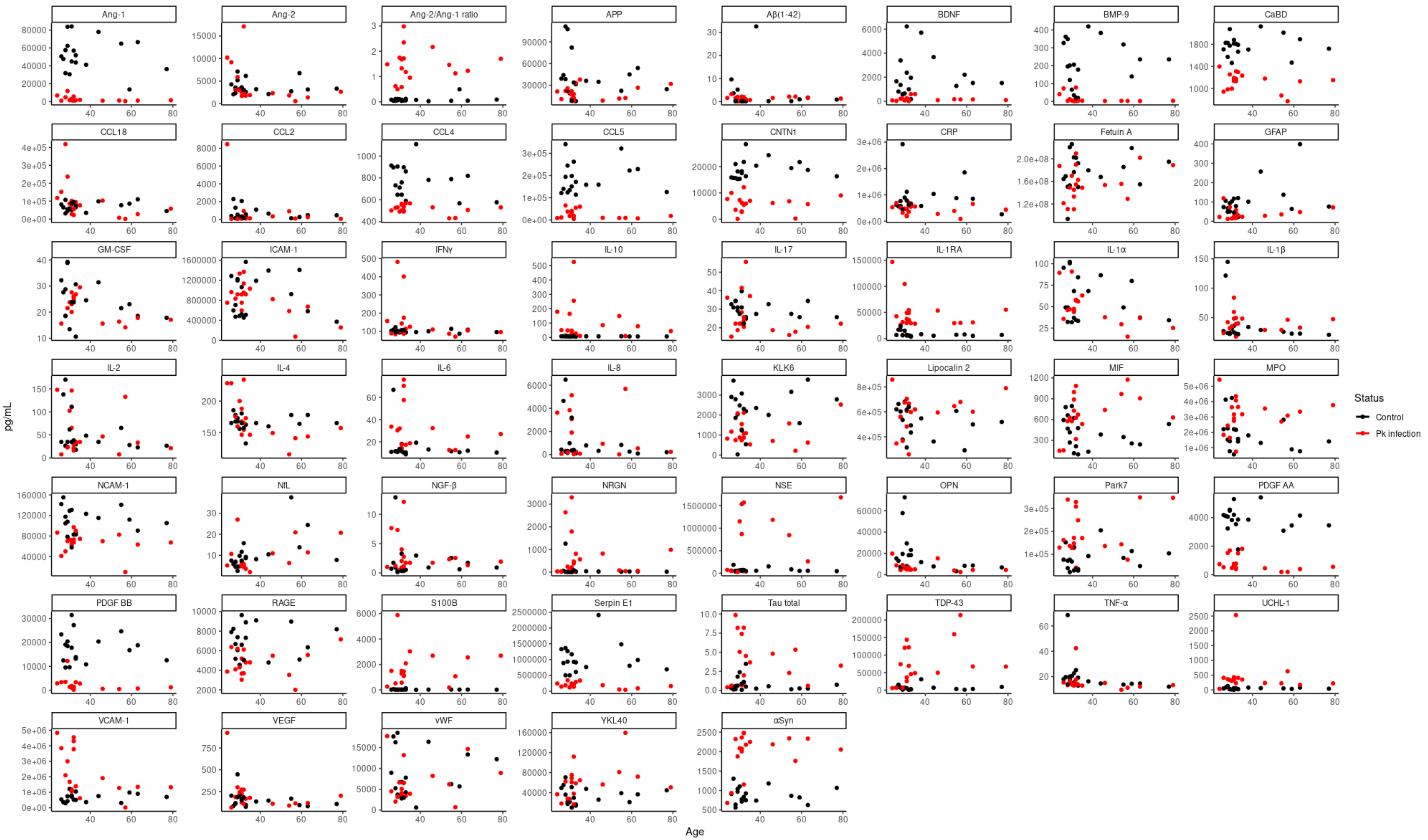

### Supplementary Figure 5

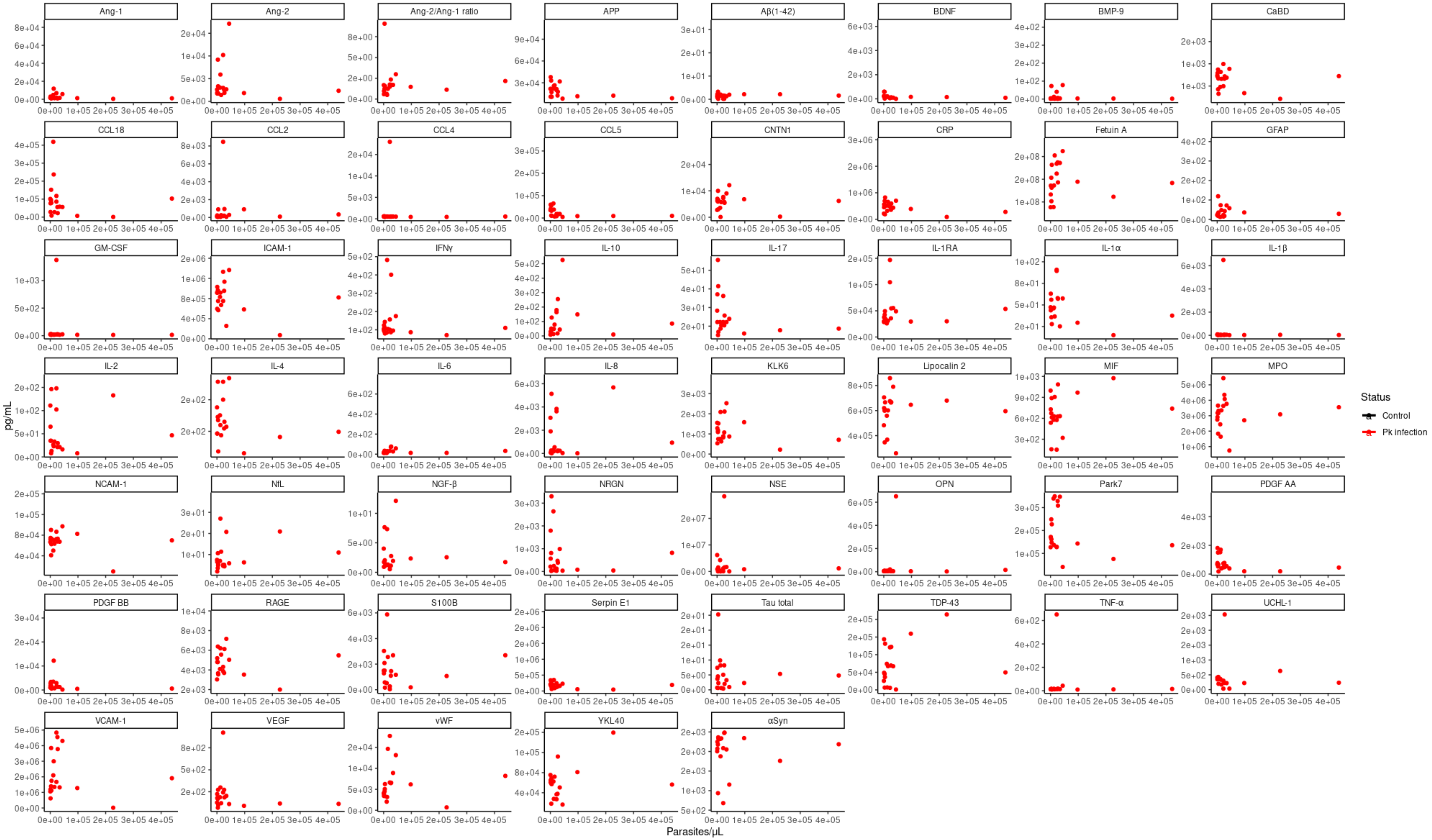

### Supplementary Figure 6

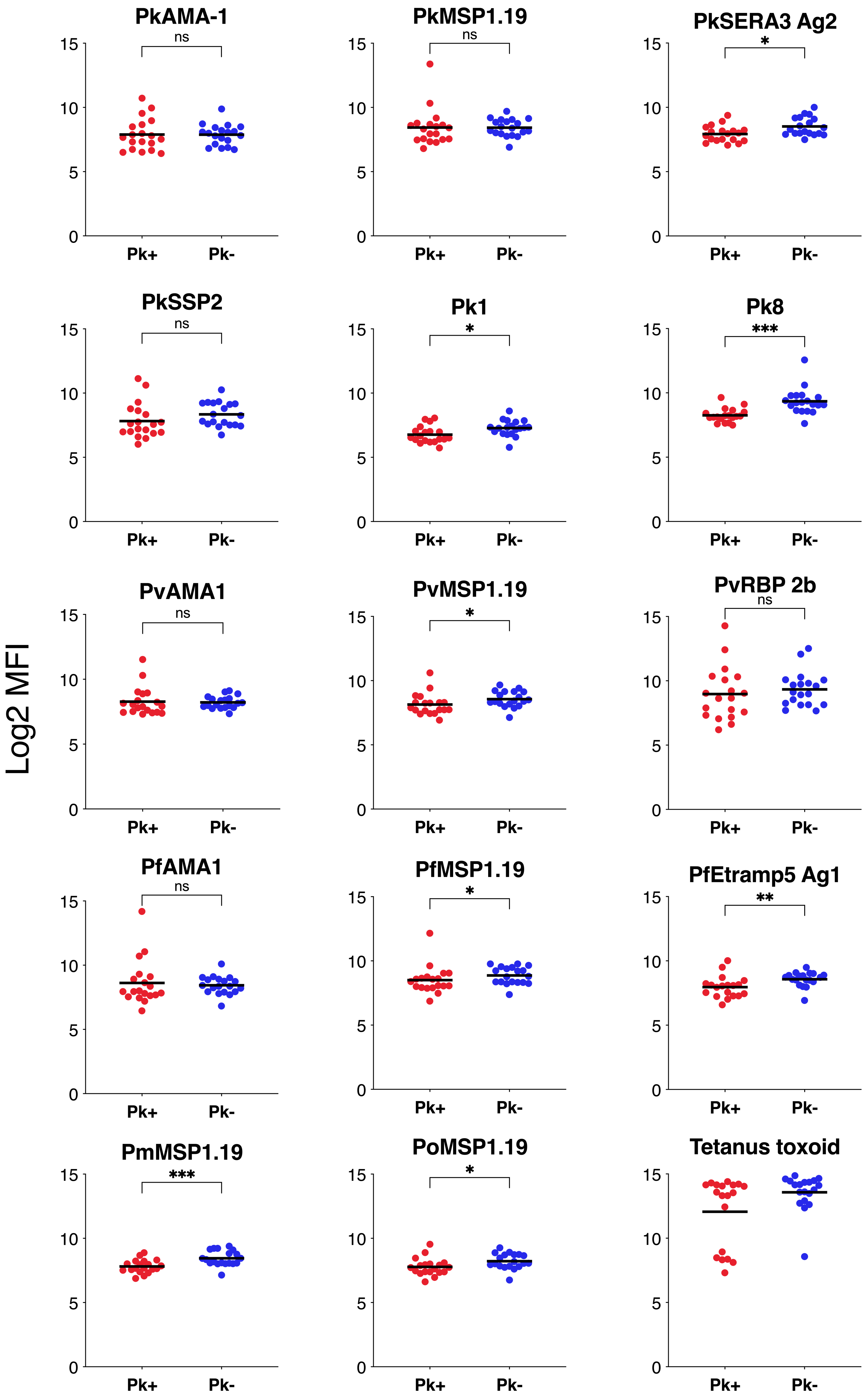

### Supplementary Figure 7

#### Serological markers of malaria exposure

Participants

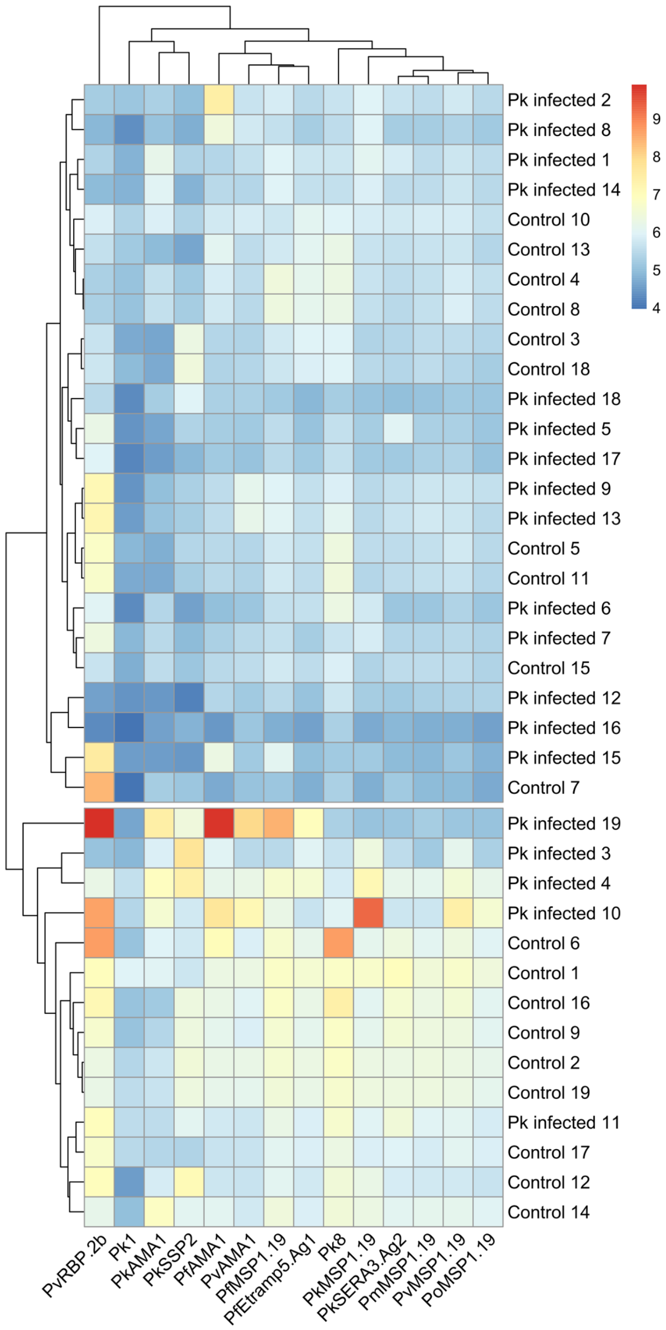
