## Supplementary Table 1 for "*Plasmodium knowlesi* infection is associated with elevated circulating biomarkers of brain injury and endothelial activation"

### Supplementary Table 1. Plasma biomarkers of infection and immune activation

#### Pro-inflammatory cytokines

| Biomarker | Clinical significance in malaria infection |
|---|---|
| **CRP** | In Ugandan children with *falciparum* cerebral malaria, plasma levels were significantly higher in retinopathy-positive patients compared with retinopathy-negative cases (1). |
| | Whole blood and plasma concentrations were significantly elevated in malaria-infected patients from Malaysia and Indonesian Papua, compared with healthy controls. Patients were mono-infected with *Plasmodium falciparum*, *vivax*, *knowlesi*, *malariae* or *ovale* as confirmed by PCR (2). |
| | In Cambodian asymptomatic participants, plasma levels were significantly higher in parasitaemic individuals compared with uninfected, age-, sex-, and village-matched controls. Patients had either a *falciparum* or *vivax* mono-infection, a *Plasmodium* infection with indeterminate species, or a mixed infection (3). |
| | In Brazilian patients with *falciparum* or *vivax* malaria, plasma levels were significantly higher in infected individuals compared with healthy controls. Levels were also higher in *vivax* patients than in *falciparum* cases (4). |
| **GM-CSF** | To the best of our knowledge, no studies have reported significant group differences in the context of human malaria. |
| **IFN-γ** | Plasma levels were significantly higher in Rwandan patients with severe malaria, compared with uncomplicated cases and controls (5). |
| | Plasma levels were significantly higher in Colombian patients with *vivax* severe malaria, compared with uncomplicated cases and healthy controls (6). |
| | In Brazilian patients with different forms of *vivax* malaria and controls, a network analysis revealed that IFN-γ, TNF-α, and CCL5 were crucial in the profile of mild malaria cases (7). |
| **IL-1α** | To the best of our knowledge, no studies have reported significant group differences in the context of human malaria. |
| **IL-1β*** | A meta-analysis revealed that IL-1β blood levels were higher in severe malaria patients compared with uncomplicated cases. *Plasmodium spp*. was a confounder in the meta-analysis, showing no difference in IL-1β levels between *falciparum*-infected groups (8). |
| | In Indian patients with *falciparum* malaria, IL-1β levels were significantly the highest in severe cases with no brain injury, compared with other groups (9). |
| | In Beninese children with *falciparum* cerebral malaria, plasma levels were significantly higher in fatal cases compared with those who survived (10). |
| **IL-2** | Compared with Mozambican adults with life-long exposure to *falciparum* malaria, Spanish travellers diagnosed with malaria had significantly higher serum IL-2 levels (11,12). |
| **IL-6*** | Plasma levels were significantly higher in Rwandan patients with severe malaria, compared with uncomplicated cases and controls (5). |
| | Plasma IL-6 levels were significantly higher in Colombian patients with *vivax* severe malaria, compared with uncomplicated cases and healthy controls (6). |
| | Plasma levels were increased in Brazilian patients with *vivax* malaria, compared with non-infected subjects with previous malaria episodes. Infected patients showed a strong correlation between CCL2 and IL-6 plasma levels, and moderate correlations between IL-6 and IL-10 (13). |
| | In Pakistani patients with *vivax* malaria, plasma levels were significantly higher in uncomplicated cases than healthy controls, and in complicated cases than in uncomplicated ones (14). |
| **IL-8** | In Beninese children with *falciparum* cerebral malaria, plasma levels were significantly higher in fatal cases compared with those who survived. In the former, IL-8 was identified as a risk factor for death by multivariate analysis (10). |
| **IL-17A** | Plasma levels were significantly higher in Ghanian children with severe *falciparum* malaria, compared with uncomplicated cases and non-malaria febrile controls (15). |
| | In Indian patients with *falciparum* malaria, plasma levels were higher in patients with multi-organ dysfunction compared with other severe malaria subgroups, suggesting IL-17 plays a role in renal inflammatory pathology during *falciparum* infection (16). |

| | |
|---|---|
| | Plasma levels were significantly higher in Rwandan patients with severe malaria, compared with uncomplicated cases and controls (5). |
| | Plasma levels were significantly higher in Brazilian patients with *vivax* malaria, compared with previously exposed, non-infected subjects and unexposed healthy donors (13). |
| **MIF** | Host MIF plasma concentrations positively correlated with *vivax* MIF levels in the plasma of Chinese patients with uncomplicated malaria (17). |
| **MPO** | In Cameroonian participants, plasma MPO levels were significantly higher in patients with *falciparum* malaria when compared with negative controls (18). |
| **TNF-α** | Serum levels were significantly higher in Indian patients with *falciparum* severe malaria and cerebral malaria, compared with healthy controls (19). |
| | In Beninese children with *falciparum* cerebral malaria, plasma levels were significantly higher in fatal cases compared with those who survived (10). |
| | TNF-α-producing monocytes were significantly lower in Malawian children with *falciparum* malaria compared with healthy controls. Cerebral malaria cases had the lowest values (20). |
| | Plasma levels were significantly higher in Rwandan patients with severe malaria, compared with uncomplicated cases and controls (5). |
| | In Brazilian patients with different forms of *vivax* malaria and controls, a network analysis revealed that IFN-γ, TNF-α, and CCL5 were crucial in the profile of mild malaria cases (7). |
| | In Pakistani patients with *vivax* malaria, plasma levels were significantly higher in complicated cases compared with uncomplicated ones. TNF-α, IL-10, ICAM-1 and VCAM-1 were the best individual predictors of complicated *vivax* malaria (14). |

#### Anti-inflammatory cytokines

| Biomarker | Clinical significance in malaria infection |
|---|---|
| **IL-1RA** | Most abundant cytokine measured in the serum of Malaysian Borneo patients with *falciparum*, *vivax*, or *knowlesi* malaria. Serum levels correlated with parasitaemia in subjects infected with any of the parasite species, and was associated with complications in *knowlesi*-infected patients (21). |
| **IL-4**** | Plasma levels were significantly higher in Colombian patients with *vivax* severe malaria, compared with uncomplicated cases and healthy controls (6). |
| | In Brazilian patients with different forms of *vivax* malaria and controls, a network analysis revealed a protective role of IL-4 and IL-10 in non-infected and asymptomatic patients (7). |
| **IL-10** | In Beninese children with *falciparum* cerebral malaria, plasma levels were significantly higher in fatal cases compared with those who survived (10). |
| | Plasma levels were significantly higher in Rwandan patients with severe malaria, compared with uncomplicated cases and controls (5). |
| | Plasma levels were significantly higher in Colombian patients with *vivax* severe malaria, compared with uncomplicated cases and healthy controls (6). |
| | IL-10 production was only observed in Brazilian patients with *vivax* malaria, compared with previously exposed, non-infected subjects and unexposed healthy donors. Infected patients presented a moderate correlation between IL-10 and IL-6 plasma levels (13). |
| | In Brazilian patients with different forms of *vivax* malaria and controls, a network analysis revealed a protective role of IL-10 and IL-4 in non-infected and asymptomatic *vivax* patients (7). |
| | In Pakistani patients with *vivax* malaria, plasma levels were significantly higher in uncomplicated cases than healthy controls, and higher again in complicated cases compared with uncomplicated ones. IL-10, TNF-α, ICAM-1 and VCAM-1 were the best individual predictors of complicated *vivax* malaria (14). |

#### Chemokines

| Biomarker | Clinical significance in malaria infection |
|---|---|
| **CCL2** | Plasma levels were significantly higher in Colombian patients with *vivax* severe malaria, compared with uncomplicated cases and healthy controls (6). |
| | Plasma levels were significantly increased in Brazilian patients with *vivax* malaria, compared with previously exposed, uninfected subjects. Infected patients showed a strong correlation between CCL2 and IL-6 plasma levels (13). |

| | | |
|---|---|---|
| **CCL4** | | Plasma levels were significantly higher in Brazilian patients with acute *falciparum* and *vivax* malaria, compared with healthy controls. During the convalescent phase, levels were higher in *falciparum* patients as compared to *vivax* cases (22). |
| **CCL5** | | Serum levels were significantly lower in Indian patients with *falciparum* severe malaria, compared with uncomplicated cases and healthy controls (23). |
| | | Plasma levels were significantly lower in Brazilian patients with *vivax* malaria, compared with previously exposed, non-infected subjects and unexposed healthy donors (13). |
| | | In Brazilian patients with different forms of *vivax* malaria and controls, a network analysis revealed that CCL5, IFN-γ, and TNF-α were crucial in the profile of mild malaria cases (7). |
| **CCL18** | | To the best of our knowledge, no studies have reported significant group differences in the context of human malaria. |

#### Other immune markers

| Biomarker | Description | Clinical significance |
|---|---|---|
| **OPN** | Extracellular matrix bone protein. It can act as a cytokine, enhancing production of IFN-γ and IL-12 and reducing production of IL-10. | Plasma concentrations in Ugandan infants with *falciparum* malaria were inversely correlated with *falciparum*-specific atypical memory B cells, suggesting that OPN could have a role in the acquisition of natural immunity against malaria (24). |
| | | Plasma levels were significantly higher in Singapore patients with vascular cognitive impairment, compared with cognitively normal controls (25). |
| **RAGE** | Cell surface pattern recognition receptor. Triggers a pro-inflammatory response. | To the best of our knowledge, no studies have reported significant group differences in the context of human malaria. |

**CCL2**: chemokine (C-C motif) ligand 2; **CCL4**: chemokine (C-C motif) ligands 4; **CCL5**: chemokine (C-C motif) ligand 5; **CCL18**: chemokine (C-C motif) ligand 18; **CNS**: central nervous system; **CRP**: C-reactive protein; **CSF**: cerebrospinal fluid; **GM-CSF**: granulocyte-macrophage colony-stimulating factor; **IFN-γ**: interferon gamma; **IL-1α**: interleukin 1 alpha; **IL-1β***: interleukin 1 beta; **IL-1RA**: interleukin 1RA; **IL-2**: interleukin 2; **IL-4****: interleukin 4; **IL-6***: interleukin 6; **IL-8**: interleukin 8; **IL-10**: interleukin 10; **IL-17A**: interleukin 17; **ILC**: innate lymphoid cells; **MIF**: migration inhibitory factor; **MPO**: myeloperoxidase; **NK**: natural killer; **OPN**: osteopontin; **RAGE**: receptor for advanced glycation end-products; **TNF-α**: tumour Necrosis Factor alpha; **VSMC**: vascular smooth muscle cells.

*May have anti-inflammatory functions. **May have pro-inflammatory functions.

**Legend**: Clinical findings related to…

*Plasmodium falciparum* malaria infection; *Plasmodium vivax* malaria infection; *Plasmodium knowlesi* malaria infection; Neurological conditions.
