## Supplementary Table 2 for "*Plasmodium knowlesi* infection is associated with elevated circulating biomarkers of brain injury and endothelial activation"

### Supplementary Table 2. Vascular biomarkers

| Biomarker | Description | Clinical significance |
|---|---|---|
| **Ang-1** | Vascular growth factor. Regulates angiogenesis, endothelial cell survival, and proliferation. Mediates blood vessel maturation and stability. | A decline in Ang-1 plasma levels was associated with increasing *falciparum* and *vivax* malaria severity and widespread endothelial activation across African and Asian patient studies, irrespective of age (1–5). |
| **Ang-2** | Vascular growth factor, biomarker of endothelial activation. Modulates Ang-1 signalling, and in the absence of angiogenic inducers such as VEGF, promotes vascular regression. In concert with VEGF, triggers a permissive angiogenic signal. Involved in lymphangiogenesis. | An increase of Ang-2 plasma levels was associated with increasing *falciparum* and *vivax* malaria severity and widespread endothelial activation across African and Asian patient studies, irrespective of age (1,3,5). |
| | | Robust predictor of mortality in *falciparum* cerebral malaria, identified as a risk factor for blood-brain barrier dysfunction, neuroinflammation, and long-term cognitive injury in African children (4,6,7). |
| $\frac{\mathbf{Ang2}}{\mathbf{Ang1}}$ | Ratio between Ang-2 and Ang-1 plasma levels. | Plasma ratio was higher in patients with *falciparum* and *vivax* severe malaria compared with uncomplicated malaria and healthy controls, across African, Asian, and Latin American patient studies and irrespective of age. Fatal cerebral malaria cases showed the highest ratio (1,2,5,8–10). |
| **BMP-9** | Growth factor, member of the TGF-β superfamily. Regulates angiogenesis by inhibiting VEGF-induced endothelial cell migration and proliferation. | To our knowledge, no studies have reported significant group differences in the context of human malaria. |
| **ICAM-1** | Intercellular adhesion molecule constitutively expressed on the vascular endothelium. Upon IL-1 and TNF-α stimulation, expression levels increase, so that leukocytes can bind to it and transmigrate into tissues. Increased plasma levels are suggestive of endothelial activation. | Plasma levels were significantly higher in Ugandan children with *falciparum* severe malaria, compared with healthy controls (4). |
| | | Plasma levels were higher in Malawian children with *falciparum* cerebral malaria and retinopathy, compared with their retinopathy-negative counterparts (11). |
| | | In Ugandan children with *falciparum* malaria, plasma levels were elevated in severe malarial anaemia fatalities compared to survivors (12). |
| | | In Ghanian children with *falciparum* cerebral malaria, plasma levels were higher compared with uncomplicated malaria cases (13). |
| | | In Pakistani patients with *vivax* malaria, plasma levels were significantly higher in uncomplicated cases than healthy controls, and higher again in complicated cases compared with uncomplicated ones. ICAM-1, VCAM-1, TNF-α and IL-10 were the best individual predictors of complicated *vivax* malaria (14). |
| **PDGF-AA** | Angiogenic promoter. Plays an important role in wound healing, and it is essential during embryonic development. | Increased plasma levels predicted abnormal cerebral blood flow and stroke in children with sickle cell disease who presented with cerebrovascular disease (15). |
| **PDGF-BB** | Angiogenic promoter. Participates in wound healing, blood vessel development, and proliferation and recruitment of pericytes and vascular smooth muscle cells in the central nervous system. | In Ugandan children with *falciparum* cerebral malaria, plasma levels were significantly higher in retinopathy-negative patients, compared to their retinopathy-positive peers (16). |
| **Serpin E1** | Serine protease inhibitor. Plays a role in the controlled degradation of blood clots. | To our knowledge, no studies have reported significant group differences in the context of human malaria. |
| **VCAM-1** | | Plasma levels were significantly higher in Ugandan children with *falciparum* severe malaria, compared with healthy controls (4). |

| | | |
|---|---|---|
| | Surface glycoprotein expressed on the vascular endothelium. Participates in immune surveillance and inflammation by regulating leukocyte adhesion to the endothelium and transendothelial migration. | In Pakistani patients with *vivax* malaria, plasma levels were significantly higher in uncomplicated cases than healthy controls, and higher again in complicated cases compared with uncomplicated ones. VCAM-1, ICAM-1, TNF-α and IL-10 were the best individual predictors of complicated *vivax* malaria (14). |
| **VEGF** | Vascular growth factor. Promotes angiogenesis, vasculogenesis, and endothelial cell growth. Induces endothelial cell proliferation, cell migration, and permeabilization of blood vessels. | Plasma levels were significantly higher in Indonesian patients with *falciparum* malaria compared with healthy controls (17). |
| | | Serum levels were significantly lower in Indian patients with *vivax* severe malaria compared with uncomplicated malaria and healthy controls (5). |
| **vWF-A2** | Glycoprotein involved in haemostasis, biomarker of endothelial activation. Promotes adhesion of platelets to sites of vascular injury and acts as a chaperone for certain coagulation factors. | Plasma levels were significantly higher in Ugandan children with *falciparum* severe malaria, compared with healthy controls (4); increased plasma levels were associated with mortality (18). |
| | | Plasma levels were significantly higher in Malawian children with cerebral malaria than in children with uncomplicated malaria, showing similar values in patients with and without retinopathy (19). |
| | | Plasma levels were significantly increased in Malaysian patients with severe and uncomplicated *vivax* malaria, compared with controls, and correlated with parasitaemia in these patients (20). |

**Ang-1**: angiopoietin-1; **Ang-2**: angiopoietin-2; **Ang-2/Ang-1**: ratio between Ang-2 and Ang-1; **BMP-9**: bone morphogenetic protein 9; **ICAM-1**: intercellular adhesion molecule 1; **PDGF-AA**: platelet-derived growth factor AA; **PDGF-BB**: platelet-derived growth factor BB; **VCAM-1**: vascular cell adhesion molecule; **Serpin E1**: Serine Proteinase Inhibitor-clade E1; **PEDF**: pigment epithelium derived factor; **VEGF**: vascular endothelial growth factor; **vWF-A2**: von Willebrand Factor (A2 domain).

**Legend**: Clinical findings related to…

*Plasmodium falciparum* malaria infection; *Plasmodium vivax* malaria infection; *Plasmodium knowlesi* malaria infection;

Neurological conditions; Other medical conditions.
