## Supplementary Table 3 for "*Plasmodium knowlesi* infection is associated with elevated circulating biomarkers of brain injury and endothelial activation"

**Supplementary Table 3. List of recombinant antigen targets utilised in the Luminex immunoassay to assess malaria exposure.**

| **Antigen name** | **Gene ID** | **Species** | **Description** | **Location** | **Marker type** |
| --- | --- | --- | --- | --- | --- |
| PkAMA1 | PKNH_0931500 | *P. knowlesi* | Apical membrane antigen 1 | Merozoite surface | Historical exposure |
| PkMSP1_19_ | PKNH_0728900 | *P. knowlesi* | Merozoite surface protein 1, 19kD | Merozoite surface | Historical exposure |
| PkSera3 Ag2 | PKNH_0413400 | *P. knowlesi* | Cysteine protease (Serine repeat-like antigen) | Parasitophorous vacuole | Utility yet to be fully characterised; evidence of short-term exposure |
| PkSSP2/TRAP | PKNH_1265400 | *P. knowlesi* | Sporozoite surface protein 2, putative, thrombospondin-related anonymous protein (TRAP) | Sporozoite surface | Utility yet to be fully characterised |
| Pk1 (PkCSP_F) | ? | *P. knowlesi* | ? | ? | Exploratory |
| Pk8 | PKNH_0400300 | *P. knowlesi* | Plasmodium exported protein, unknown function | ? | Exploratory |
| PvAMA1 | PVX_092275 | *P. vivax* | Apical membrane antigen 1 | Merozoite surface | Historical exposure |
| PvMSP1_19_ | PVX_099980 | *P. vivax* | Merozoite surface protein ,1, 19kD | Merozoite surface | Historical exposure |
| PvRBP2b | PVX_094255 | *P. vivax* | Reticulocyte binding protein 2b fragment | Merozoite micronemes | Recent exposure |
| PfAMA1 | PF3D7_1133400 | *P. falciparum* | Apical membrane antigen 1 | Sporozoite / merozoite surface | Historical exposure |
| PfMSP1_19_ | PF3D7_0930300 | *P. falciparum* | Merozoite surface protein 1, 19kD | Merozoite surface | Historical exposure |
| Etramp5 Ag1 | PF3D7_0532100 | *P. falciparum* | Early transcribed membrane protein 5 antigen 1 | Infected erythrocyte / parasitophorous vacuole membrane | Recent exposure |
| PoMSP1_19_ | PocGH01_07037900 | *P. ovale* | Merozoite surface protein 1, 19kD | Merozoite surface | Historical exposure |
| PmMSP1_19_ | PmUG01_07042000 | *P. malariae* | Merozoite surface protein 1, 19kD | Merozoite surface | Historical exposure |
| Tetanus toxoid | -- | *Clostridium tetani* | Inactivated tetanus toxin immunisation antigen; internal human control | -- | -- |
| GST | GST26_SCHJA | *Schistosoma japonicum* | Gluthanoid-S-transferase purification tag; GST-tagged protein control | -- | -- |
