## Supplementary Table 4 for "*Plasmodium knowlesi* infection is associated with elevated circulating biomarkers of brain injury and endothelial activation"

### Supplementary Table 3

##### Individual biomarker group comparisons (Wilcoxon test)

| | | **Healthy controls** (N=19) Median pg/mL (IQR) | ***Pk*-infected patients** (N=19) Median pg/mL (IQR) | **Crude P value** | **Corrected* P value** |
|---|---|---|---|---|---|
| | **αSyn** | 863.4 (259.9) | 2,177.8 (392.4) | **<0.0001** | **<0.0001** |
| | **APP** | 36,082.0 (25,945.0) | 17,701 (12,728.0) | **0.0010** | 0.05 |
| Biomarkers of brain alterations or cerebral injury | **Aβ$_{(1-42)}$** | 0.2 (1.2) | 1.6 (0.9) | 0.08 | 1.00 |
| | **BDNF** | 1,516.2 (1,454.1) | 146.2 (115.8) | **<0.0001** | **<0.0001** |
| | **CaBD** | 1,807.0 (179.0) | 1,151.1 (221.0) | **<0.0001** | **<0.0001** |
| | **CNTN1** | 18,191.0 (4,890.0) | 6,179.7 (2,615.6) | **<0.0001** | **<0.0001** |
| | **ENO2/NSE** | 54,324.0 (21,601.0) | 1,094,366.0 (1,382,298.0) | **0.0010** | 0.05 |
| | **Fetuin A** | 176,390,803.0 (38,746,703.0) | 154,460,943.0 (47,209,645.0) | **0.0462** | 1.00 |
| | **GFAP** | 78.1 (45.8) | 29.9 (25.3) | **<0.0001** | **0.0013** |
| | **KLK6** | 2,319.5 (915.0) | 1,072.4 (788.7) | **0.0003** | **0.0126** |
| | **Lipocalin 2** | 547,608.0 (149,670.0) | 619,695.0 (94,135.0) | 0.05 | 1.00 |
| | **NCAM-1** | 107,813.0 (38,204.0) | 70,203.0 (8,851.0) | **<0.0001** | **<0.0001** |
| | **NfL** | 8.1 (4.4) | 6.3 (5.8) | 0.21 | 1.00 |
| | **NGF-β** | 0.9 (1.2) | 1.7 (1.5) | **0.0203** | 1.00 |
| | **NRGN** | 22.4 (11.1) | 241.4 (763.1) | **<0.0001** | **0.0022** |
| | **Park7** | 71,436.0 (61,372.0) | 161,810.0 (146,171.0) | **<0.0001** | **0.0006** |
| | **S100B** | 27.3 (0.0) | 1,282.2 (1,777.2) | **<0.0001** | **<0.0001** |
| | **Tau total** | 0.6 (0.6) | 3. 7 (5.0) | **<0.0001** | **0.0007** |
| | **TDP-43** | 5,344.3 (5,806.0) | 67,143.0 (90,407.0) | **<0.0001** | **0.0050** |
| | **UCHL-1** | 49.3 (45. 4) | 336.6 (184.1) | **<0.0001** | **<0.0001** |
| | **YKL40** | 29,591.0 (24,732.0) | 58,674.0 (33,187.0) | **0.0020** | 0.10 |

*Bonferroni correction for multiple comparisons.

| Category | Biomarker | **Healthy controls** (N=19) Median pg/mL (IQR) | ***Pk*-infected patients** (N=19) Median pg/mL (IQR) | **Crude P value** | **Corrected* P value** |
|---|---|---|---|---|---|
| Pro-inflammatory cytokines | **CRP** | 668,496.0 (308,499.0) | 477,838.0 (235,914.0) | **0.0005** | **0.0280** |
| | **GM-CSF** | 23.8 (10.0) | 21.52 (8.7) | 0.24 | 1.00 |
| | **IFNγ** | 101.4 (11.8) | 104.8 (46.3) | 0.73 | 1.00 |
| | **IL-1α** | 49.0 (46.4) | 46.0 (21.2) | 0.29 | 1.00 |
| | **IL-1β** | 24.7 (7.2) | 40.0 (17.9) | **0.0027** | 0.14 |
| | **IL-2** | 34.8 (28.7) | 33.1 (62.5) | 0.53 | 1.00 |
| | **IL-6** | 11.5 (2.6) | 21.7 (19.2) | **0.0013** | 0.06 |
| | **IL-8** | 319.7 (599.0) | 254.6 (2,353.2) | 0.82 | 1.00 |
| | **IL-17A** | 29.2 (6.1) | 22.2 (8.3) | **0.0081** | 0.41 |
| | **MIF** | 384.6 (315.8) | 630.7 (321.2) | **0.0022** | 0.11 |
| | **MPO** | 1,571,073.0 (869,998.0) | 3,176,104.0 (919,812.0) | **0.0006** | **0.0314** |
| | **TNF-α** | 16.2 (5.7) | 13.7 (2.3) | **0.0393** | 1.00 |
| Anti-inflammatory | **IL-1RA** | 6,736.0 (2,506.0) | 35,639.0 (21,569.0) | **<0.0001** | **<0.0001** |
| | **IL-4** | 165.1 (12.2) | 159.8 (35.1) | 0.44 | 1.00 |
| | **IL-10** | 8.0 (1.3) | 48.6 (116.0) | **<0.0001** | **<0.0001** |
| Chemokines | **CCL2** | 380.8 (326.0) | 160.9 (219.4) | **0.0075** | 0.38 |
| | **CCL4** | 781.8 (250.0) | 526.3 (47.7) | **<0.0001** | **0.0002** |
| | **CCL5** | 158,730.0 (96,050.0) | 17,650.0 (27,045.0) | **<0.0001** | **<0.0001** |
| | **CCL18** | 76,800.0 (37,565.0) | 76,703.9 (73,970.5) | 0.89 | 1.00 |
| Other | **OPN** | 12,981.0 (11,051.0) | 5,057.0 (3,291.0) | **0.0033** | 0.17 |
| | **RAGE** | 7,206.0 (2,599.0) | 4,797.0 (1,840.0) | **<0.0001** | **0.0025** |

**Biomarkers of infection and immune activation**

*Bonferroni correction for multiple comparisons.

| | | **Healthy controls** (N=19) Median pg/mL (IQR) | ***Pk*-infected patients** (N=19) Median pg/mL (IQR) | **Crude P value** | **Corrected* P value** |
|---|---|---|---|---|---|
| Vascular biomarkers | **Ang-1** | 51,807.0 (23,226.0) | 1,547.8 (1,633.8) | **<0.0001** | **<0.0001** |
| | **Ang-2** | 3,187.0 (1,277.0) | 2,561.7 (1,365.2) | **0.0471** | 1.00 |
| | **Ang-2/Ang-1 ratio** | 0.1 (0.1) | 1. 5 (0.6) | **<0.0001** | **<0.0001** |
| | **BMP-9** | 205.9 (193.2) | 3.6 (2.3) | **<0.0001** | **<0.0001** |
| | **ICAM-1** | 705,782.0 (710,688.0) | 832,537.0 (324,397.0) | 0.98 | 1.00 |
| | **PDGF AA** | 3,864.0 (689.0) | 552.6 (346.2) | **<0.0001** | **<0.0001** |
| | **PDGF BB** | 18,495.0 (7,646.0) | 1,385.7 (2,055.8) | **<0.0001** | **<0.0001** |
| | **Serpin E1** | 932,450.0 (468,184.0) | 183,488.0 (103,411.0) | **<0.0001** | **<0.0001** |
| | **VCAM-1** | 698,022.0 (419,633.0) | 1,673,430.0 (2,154,222.0) | **<0.0001** | **0.0001** |
| | **VEGF** | 147.3 (81.2) | 178.9 (118.7) | 0.26 | 1.00 |
| | **vWF** | 6,716.8 (8,770.7) | 6,173.0 (3,737.2) | 0.20 | 1.00 |
